# Disease-economy trade-offs under alternative pandemic control strategies

**DOI:** 10.1101/2021.02.12.21251599

**Authors:** Thomas Ash, Antonio M. Bento, Daniel Kaffine, Akhil Rao, Ana I. Bento

**Author notes:** We thank Jude Bayham, Matthew G. Burgess, Dana Goldman and Adam Rose for helpful comments.

## Abstract

Public policy and academic debates regarding pandemic control strategies note potential disease-economy trade-offs, and often prioritize one outcome over the other. Using a calibrated, coupled epi-economic model of individual behavior embedded within the broader economy during a novel epidemic, we show that targeted isolation strategies can avert up to 91% of individual economic losses relative to voluntary isolation strategies. Notably, the economic savings from targeted isolation strategies do not impose an additional disease burden, avoiding disease-economy trade-offs. In contrast, widely-used blanket lock-downs do create sharp disease-economy trade-offs and impose substantial economic costs per additional case avoided. These results highlight the benefits of targeted isolation strategies for disease control, as targeted isolation addresses the fundamental coordination failure between infectious and susceptible individuals that drives the recession. Our coupled-systems framework uses a data-driven approach to map economic activities to contacts, which facilitates developing effective control strategies for future novel pathogens. Implementation of this framework can help control disease spread and potentially avert trillions of dollars in losses.

## Introduction

To date, over 106 million individuals have been infected with SARS-CoV-2 and more than 2.3 million have died worldwide, with over twenty percent of these deaths happening in the United States [1]. The pandemic also triggered the sharpest economic recession in modern American history. According to the Commerce Department, during the second quarter of 2020 US Gross Domestic Product shrank at an annual rate of 32.9 percent [2]. The COVID-19 pandemic’s rapid growth exposed a need for coupled-systems frameworks that link epidemiological and economic models and assess potential disease-economy trade-offs. Such frameworks can reveal important features of control strategies, such as the role of targeted isolation strategies that can overcome the fundamental coordination failure between infectious and susceptible individuals that drives the economic recession.

Broadly, four areas of study have informed the assessment of control strategies. Epidemiological studies evaluate disease dynamics and consider the heterogeneity of impacts resulting from control strategies [3, 4, 5, 6, 7, 8, 9, 10, 11, 12]. Epi-economics studies consider the micro-foundations of human behavior as drivers of the disease, as well as the costs and benefits of alternative control strategies [13, 14, 15, 16, 17, 18, 19, 20, 21, 22, 23]. An emerging literature on the macroeconomic consequences of pandemics considers the impacts of COVID-19 and various control strategies, either by embedding these behaviors in a broader economy with disease dynamics [24, 25] or by conducting detailed macroeconomic projections without disease dynamics [26, 27]. In addition, numerous statistical analyses have examined the relationship between disease-related behaviors and economic activity [28, 29, 30]. Several knowledge gaps remain. For example, structurally mapping economic activities to contacts in a tractable fashion that retains the underlying heterogeneity of the population presents various challenges. One major challenge is how to calibrate this mapping using epidemiological social contact surveys, which contain data on potentially disease-transmitting contacts between individuals. Further, detailed individual behavior and epidemiological transmission mechanisms have typically not been embedded into models that consider the broader economy. Finally, the set of control strategies considered in coupled-systems models remains limited and overly-simplified and, to date, have not included individual-focused targeted isolation strategies that could overcome disease-economy trade-offs.

To address these gaps, we develop a coupled epi-economic model of individual behavior embedded within the broader economy. Fig.1 presents a schematic representation of our model. Individuals make consumption and labor-leisure choices, considering the risk of infection. These decisions generate individual contacts which evolve endogenously in the model—i.e., contact rates affect and are affected by the disease dynamics. Depending on the activity, contacts can be avoidable or unavoidable. For example, given the structure of the U.S. economy, the average individual has around 7.5 contacts at their place of work during an 8-hour workday. These are avoidable contacts if the individual can alter their labor supply. In contrast, contacts such as those that occur at home are unavoidable, albeit not devoid of risk of infection [31]. Infection causes a productivity loss, as only individuals with no or mild symptoms will be able to work. The model solves the individual’s dynamic optimization problem of balancing risk and activity and aggregates the solution across the population, permitting direct calculation of disease-economy trade-offs. Through contacts, infectious individuals put susceptible individuals at risk, and in the absence of control strategies, bear no direct consequences for this imposition (in economics terminology, an infectious individual creates a *negative externality* on a susceptible individual). As the disease progresses, susceptible individuals engage in averting behavior by cutting consumption and labor, contributing to the economic recession. For analytical tractability, the framework assumes individuals have full information about their health status, although the presence of pre-symptomatic and asymptomatic individuals is reflected in the calibration of productivity losses (see SI 2.1.1).

**Figure 1:**
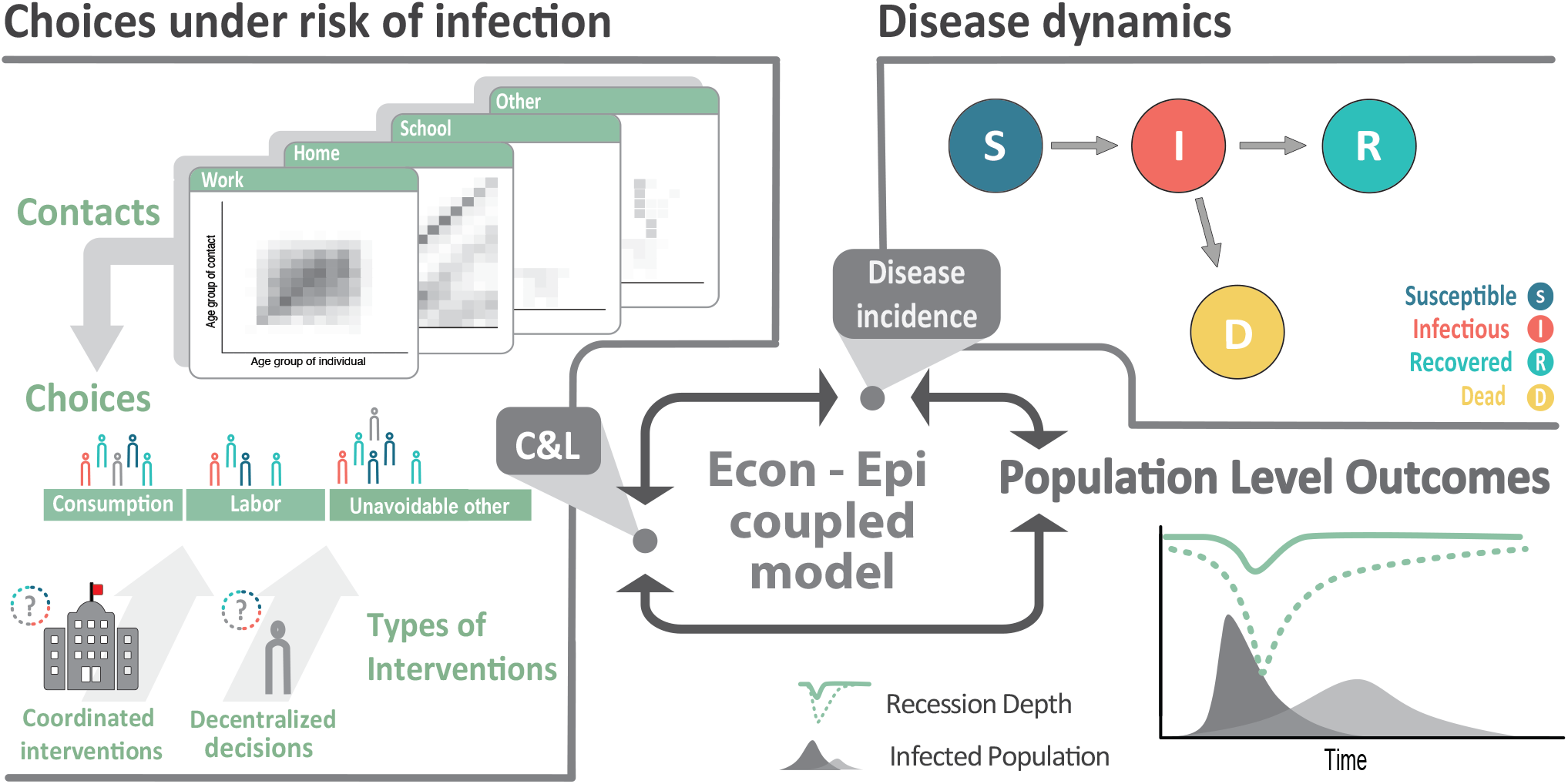
Coupled system schematic. Individuals make consumption and labor-leisure choices, considering the risk of infection through contacts with others. Individual choices and resulting contacts affect and are affected by the disease dynamics. Individual economic choices drive population-level outcomes such as disease prevalence and economic recessions. Under decentralized approaches, individuals optimize their behaviors based on their own preferences and health status. Under coordinated approaches, individuals’ behaviors are optimized based on how they affect population-level outcomes.

A key challenge for disease control and avoiding economic losses is the inability of susceptible individuals to coordinate with infectious individuals and encourage them to reduce their activities and contacts [32]. Absent such coordination, susceptible individuals bear the full burden of adjusting their consumption and labor to minimize personal risk. In a world where this coordination failure is solved, e.g., by paying infectious individuals to isolate, susceptible individuals could still consume, work, and engage in contacts, minimizing individual economic losses and the resulting recession. Thus, there is a need for control strategies that target infectious individuals. Importantly, to illustrate the benefits of such targeting strategies, we abstract from many aspects of individual heterogeneity, often captured in epidemiological studies (e.g., [33, 34, 9, 35]). This is reasonable, since the fundamental coordination failure is itself independent of heterogeneity (see SI 2.5).

We calibrate our model to pre-pandemic economic and social mixing data, using 2017 contact survey data from [6] and next-generation matrix methods [36] to generate a contact function linking different economic activities to contacts and to calibrate the transmission rate (see SI 2.3.1 and 2.3.3 for the contact matrices used and an overview of next-generation method). We study the disease-economy trade-offs that result from three alternative control strategies: voluntary isolation, blanket lockdown, and targeted isolation. Under voluntary isolation, individuals continue to optimize their personal behavior based on preferences and health status. Some may isolate, others will not. With a blanket lockdown, all individuals are forced to isolate, independent of their disease status. In the U.S., for example, by April 15 2020, more than 95% of the population was under a stay-at-home order [37, 29]; such social distancing policies come with very large economic costs and social disruption. Finally, targeted isolation of infectious individuals is able to address the coordination failure, effectively separating susceptible individuals from infectious ones (see Materials and Methods).

Implementing targeted isolation strategies comes with challenges. While our focus is on the benefits that targeted isolation strategies have the potential to deliver, we also sketch out some of the key implementation details in the discussion. These include access to rapid testing, likely heterogeneity in contacts across individuals, and limited information in the early stages of the epidemic. Focusing on targeted isolation under unconstrained access to rapid testing allows us to highlight the coordination failure that drives disease-economy trade-offs.

We show this lack of coordination is economically costly: widely-used control strategies, such as voluntary isolation or blanket lockdowns suppress the epidemic nearly as effectively as targeted isolation, but impose a much deeper recession. Targeted isolation strategies avoid these sharp disease-economy trade-offs by incentivizing infectious individuals to isolate. This allows susceptible individuals to continue to consume and work, carrying the economy through the epidemic with a milder recession. Using targeted isolation strategies instead of voluntary isolation strategies can avert substantial costs—on the order of $3.5 trillion in averted recessionary losses.

## Results

We highlight two main findings. First, regardless of control strategy, in our model the SARS-CoV-2 epidemic spreads rapidly in the population, with peak daily incidence early in the epidemic (Fig.2A & C), and final proportions of the population exposed (Fig.2C) and fatalities (Fig. S2) are largely unaltered. Second, under a targeted isolation strategy, disease control does not come at as large an economic cost as under the two other control strategies. (Fig.2).

**Figure 2:**
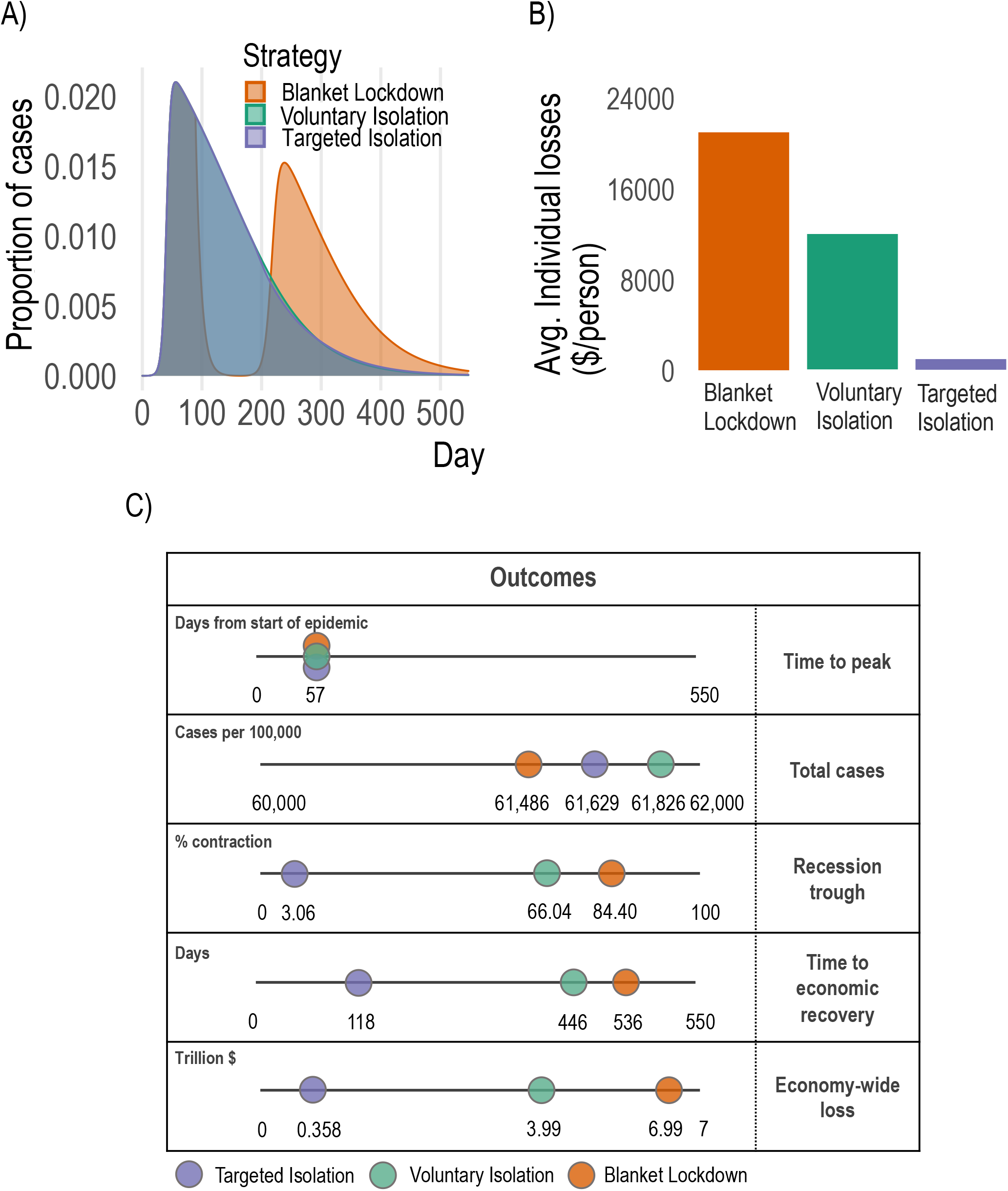
Disease dynamics and economic outcomes under voluntary isolation, blanket lockdown, and targeted isolation. **A:** Proportion of population infected over time under each strategy. Voluntary isolation and targeted isolation curves are almost-entirely overlapping, indicating nearly-identical disease dynamics. **B:** Individual losses incurred under each strategy. **C:** Key aggregate disease and economy outcomes under each strategy.

Targeted isolation strategies optimally coordinates individuals’ behavior over the course of the epidemic (Fig.2B), reducing average individual consumption losses from around $12,078 under an uncoordinated, voluntary isolation strategy to around $1,082. Targeted isolation thus delivers a 91% reduction in losses experienced by an average individual (Fig.2C) who earns $58,000 per year in the pre-epidemic baseline (Table S2). In aggregate terms, this is the difference between a peak-to-trough economic contraction of around 66% under voluntary isolation (an historically-severe recession; in our model the *quarterly* peak-to-trough drop is 33% compared with the actual drop in US quarterly consumption of 38%, see SI section 5.5) and a peak economic contraction of around 3% under targeted isolation (a mild and not-atypical recession).

Moreover, targeted isolation also confers significant benefits over blanket lockdown approaches: a targeted isolation strategy can avert approximately 95% of the recessionary losses under the case-minimizing blanket lockdown (a 75-day blanket lockdown applied to 85% of the population beginning at day 91 of the epidemic spread; see SI 4.1). This blanket lockdown only averts around 0.2% of total cases incurred under a targeted isolation policy (Fig.2). This result is robust to an empirically-plausible range of blanket lockdown designs (Fig. S4). Targeted isolation strategies reduce the depth of the recession and the duration as well (Fig.2). Fig.2 shows recovery to 1.5% of pre-epidemic GDP would occur in 118 days under a targeted isolation strategy, compared to 536 days under a blanket lockdown. While implementing the blanket lockdown reduces cases, it leads to a rebound (Fig.2A) when the lockdown is relaxed (observed in all blanket lockdown scenarios considered—Fig. S4 & Fig. S7).

The large economic savings (averted losses) and the marked difference in the probability of contact (Fig.3B) from the targeted isolation strategy arise primarily from shifting the burden of isolating from susceptible to infectious individuals (Fig.3A). Under a voluntary isolation strategy, some infectious individuals continue to work and consume despite the risk they impose on others [38, 39, 40]. This is the key coordination failure that increases the probability of infection (Fig.3B) and forces susceptible individuals to work and consume less to avoid infection (Fig.3A). Since susceptible individuals are the majority of the population in a novel epidemic, this approach to disease control comes at a large economic cost. By contrast, targeting isolation at infectious individuals dramatically changes the composition of the pool of people working and consuming (Fig.3A & B). At an individual level, voluntary isolation at the epidemic peak leads to about 3 fewer hours spent at consumption activities and 6 fewer hours spent at labor activities per day, while targeted isolation imposes similar adjustments, but just on infectious individuals (Table S2). This does not cause changes in mean daily contacts between strategies (Fig.3C & D), nor prevalence by activity type (Fig.3E), even though many more susceptible individuals are able to work and consume. As a consequence, targeting delivers small improvements in infection outcomes but massive economic savings.

**Figure 3:**
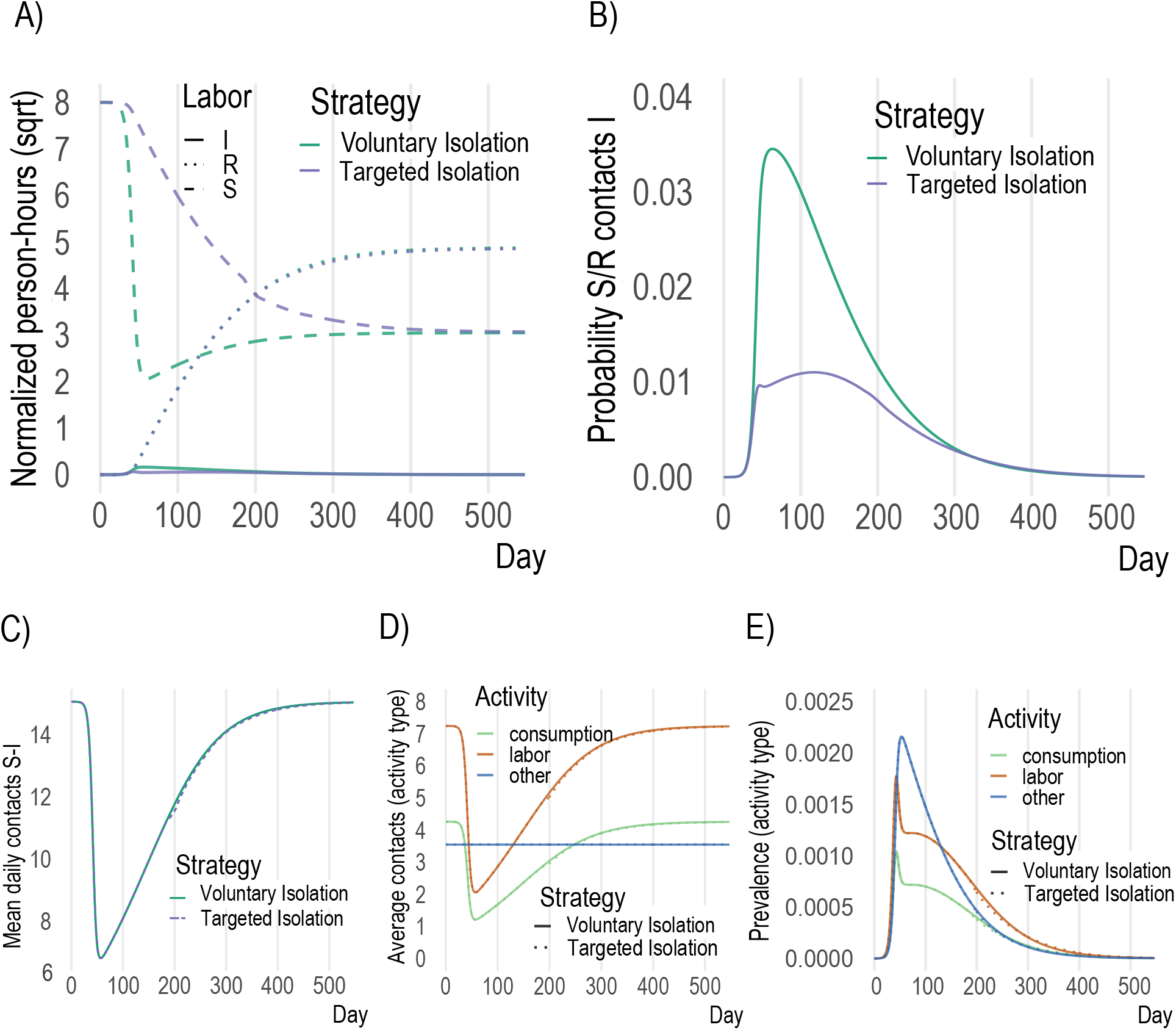
Key model mechanisms. **A**: in voluntary isolation susceptible individuals withdraw from activity due to the presence of infectious individuals (green dashed), while under targeted isolation susceptible agents engage in much more activity (blue dashed). **B**: more infectious agents at activity sites under voluntary isolation leads to higher probability of infection throughout epidemic. **C**,**D**,**E**: overall contacts, contacts by activity and prevalence (% infectious) do not change meaningfully across voluntary and targeted isolation, as the same infection outcomes are achieved despite enabling far more activity by susceptible individuals with targeted isolation.

One might be concerned about the robustness of our conclusions to simplifications and modeling choices, particularly the impact of asymptomatic individuals, our use of pre-pandemic contact data to calibrate the contact function, and the implication (through the functional form of the contact function) that individuals have identical contact levels. First, while we do not explicitly model a compartment for asymptomatic individuals, we implicitly account for them through productivity losses as shown in Fig.4A. Second, because structural changes in the economy during the pandemic may have reduced the number of contacts per unit of activity (e.g., increased prevalence of contactless goods delivery), we examine our findings’ robustness by altering the ratios of contacts at different activities and changing the functional form of the contact function. Fig.4B-E show how our conclusions about targeted isolation relative to voluntary isolation change as we vary the contact structure of the economy and the proportion of non-severely-diseased individuals. From the white dots (main model calibration), moving to the left in Fig.4B-E shows that lower prevalence of asymptomatic individuals will increase the economic effectiveness of targeted isolation without affecting the relative number of cases averted. Moving up the vertical axis in Figs.4B-C shows that increasing the share of contacts which occur at consumption rather than labor activities (e.g., if remote work becomes more common while bars and restaurants remain open) would again increase the economic effectiveness of targeted isolation without affecting the relative number of cases averted. Moving up the vertical axis of Fig.4D-E shows that increasing the share of contacts at unavoidable activities (e.g., if consumption and labor become increasingly contactless) will reduce the economic effectiveness of targeted isolation without affecting the relative number of cases averted. The functional form of the contact function allows us to examine heterogeneity in contact rates—Fig.4F-G. Convex contact functions emerge when high-contact activities (individuals) are reduced (isolated) first and concave functions emerge when high-contact activities (individuals) are reduced (isolated) last. Targeted isolation accounting for these choices is likely to produce convex contact functions if high-contact activities (individuals) are reduced (isolated) first. We find such variations have a modest impact on the economic effectiveness of targeted isolation, but do not affect its disease control properties. We discuss these forms further in Materials and Methods and the SI.

**Figure 4:**
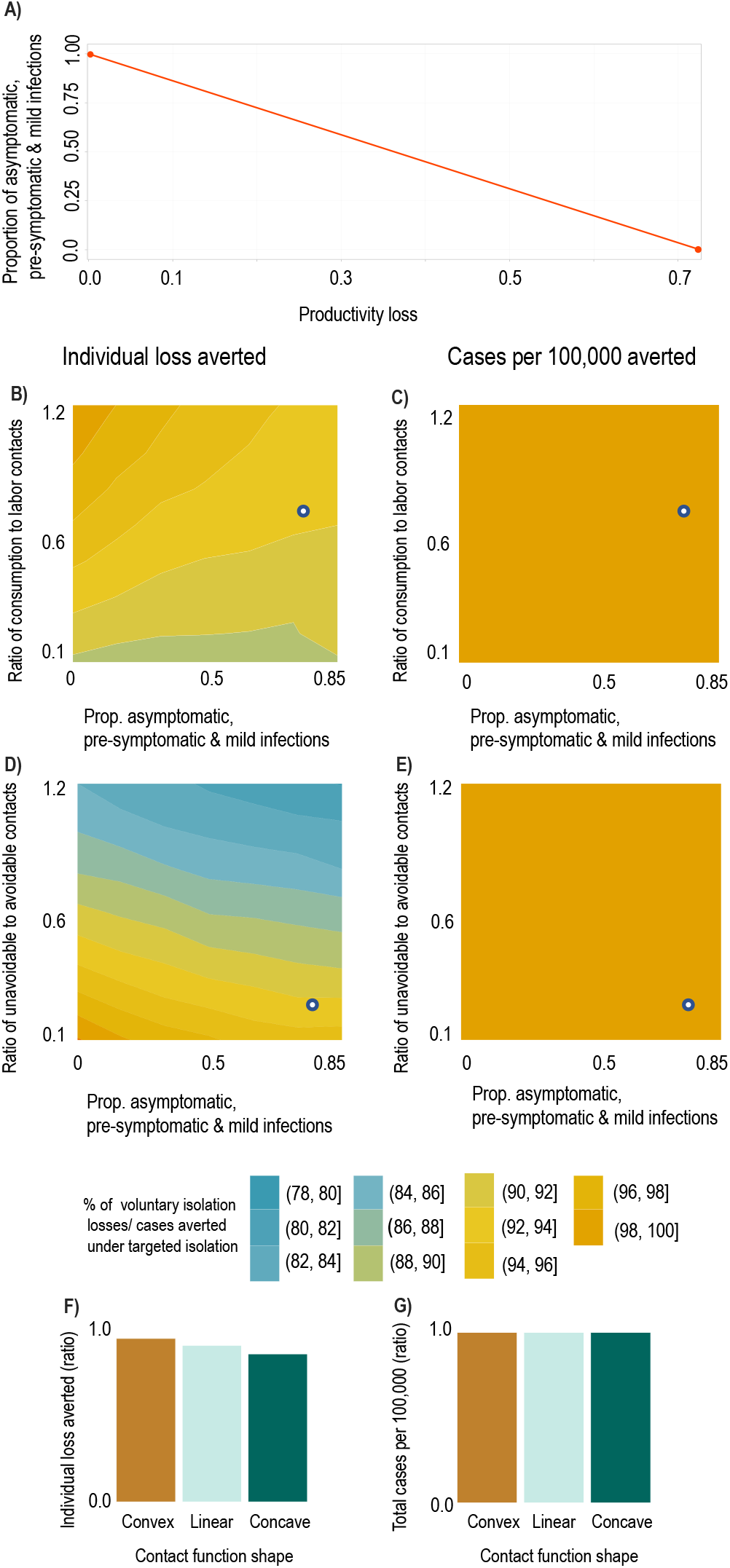
Result sensitivity to key model parameters. We plot ratios of outcomes under targeted vs. voluntary isolation to highlight the relative variation in outcomes under each strategy. The white dots in panels A and C show the baseline parameterization. **A:** Mapping between productivity losses and implied share of the population which is pre-symptomatic, asymptomatic, or has mild symptoms (i.e., infectious individuals able to work). A productivity loss of 0.85 implies approximately 80% of the population are pre-symptomatic, asymptomatic, or have mild symptoms. **B, C:** Ratio of individual losses averted (B) and ratio of cases per 100k averted (C) under targeted isolation vs. voluntary isolation as proportion of contacts at consumption relative to labor activities increases (a value of 1 means equal number of contacts at consumption and labor) and as the asymptomatic share increases. **D, E:** Ratio of individual losses averted (D) and ratio of cases per 100k averted (E) under targeted isolation vs. voluntary isolation as proportion of unavoidable contacts (e.g., home) relative to avoidable contacts (consumption & labor) increases (a value of 1 means an equal number of contacts at home as at consumption & labor) and as the asymptomatic share increases. **F, G:** Ratio of individual losses averted (F) and ratio of cases per 100k averted (G) under targeted isolation vs. voluntary isolation as contact functional form varies. Convex contact functions imply high-contact activities are easiest to avoid, while concave contact functions imply low-contact activities are easiest to avoid (see Materials and Methods).

## Discussion

A year into the SARS-CoV-2 pandemic, it is increasingly clear economic concerns cannot be neglected [41]. We show a targeted isolation approach emerges from our model as an optimal strategy, balancing disease spread and economic activities. While our model simplifies several important features specific to the COVID-19 pandemic, our predicted infection rates and economic responses are broadly consistent with observed patterns, and thus our results likely capture the correct order of magnitude and capture the key qualitative features of the epidemic and recession.

Recent studies suggest the COVID-19 recession was driven by voluntary reductions in consumption in response to increasing infection risk [28, 42]. We show this drop in consumption is driven by a coordination failure: infectious individuals do not face the full social costs of their activities, leading susceptible individuals to withdraw from economic activity. This coordination failure resembles the classical problem of the tragedy of the commons in natural resources and the environment [43, 44, 45], underscoring the lack of property rights in the market for infection-free common spaces. It also shares similarities with coordination issues that emerge in climate change, fisheries, orbit use and other settings [46, 47, 48, 49]. Correcting this coordination failure via a targeted isolation strategy that internalizes the costs infectious individuals impose on susceptible individuals delivers substantial economic savings (Fig.2 & Fig.3).

Our conclusions arise from a data-driven method to calibrate the mapping between disease-transmitting contacts and economic activities. Compartmental models of infectious diseases typically segment activities based on population characteristics like age and student status (e.g., [33, 34, 7, 50, 51]) rather than economic choices like consumption and labor. We build on prior work in this area (e.g. [15, 16, 18]) to address two long-standing challenges: appropriately converting units of disease-transmitting contacts into units of economic activities (contacts into dollars and hours), and calibrating the resulting contact function to produce the desired *ℛ*_0_. We address these challenges in three steps (see Materials and Methods and SI).

First, we use contact matrices from [6] to construct age-structured contact matrices at consumption, labor, and unavoidable other activities (Fig. S1). We then use next-generation matrix methods to calculate the mean number of contacts, adjusted for how individuals of different ages mix with each other, at each activity. Finally, we use these values with pre-epidemic consumption and labor supply levels to map contacts to dollars and hours in the contact function, and then calibrate the *ℛ*_0_. This approach provides a behaviorally-grounded perspective on why contacts occur. Understanding the structure and benefits of targeted isolation requires this mapping between economic activities and contacts.

Our results also serve to highlight an important benefit of making rapid testing for novel pathogens widely available [52, 53, 54, 55]—it facilitates targeted isolation approaches which reduces economic losses. Additionally, a targeted isolation strategy may even incentivize individuals to acquire knowledge of their disease status and isolate. For example, if individuals (particularly those unable to work remotely) are compensated for income lost due to isolation, the financial cost of isolation due to a positive test is reduced. In that sense, solving the coordination problem induces a solution to the problem of unknown disease status, provided sufficient rapid tests are available.

However, we stress the fundamental problem is a missing market for infection-free common spaces, not missing information about health status. If property rights over infection-free common spaces are well-defined, susceptible individuals could pay infectious individuals to remain away from consumption and labor activities [56]. Funding to overcome the limited availability of testing is therefore akin to a transaction cost—and while transaction costs limit the efficiency of rights-based solutions [57], they do not overturn the key insight that addressing the coordination failure between susceptible and infectious individuals yields major benefits.

To implement targeted isolation, governments can provide incentives and encouragement for infectious individuals to remove themselves from these public spaces. In our model, paying individuals to stay home while infectious would require spending on the order of $428 billion (two weeks pay times the total number of infected), to purchase the gain of an avoided recession on the order of $4 trillion— total savings of $3.5 trillion relative to voluntary isolation, not including additional averted costs from long-term negative public health outcomes [27]. Our focus here is on the benefits of targeted isolation strategies rather than the details of how to implement them, or on cross-regional comparisons of implemented strategies (see SI 5.4.1 for more discussion); designing such incentive mechanisms presents its own challenges, e.g., [58, 14, 10], and is an important area for future research.

Given the appealing features of targeted isolation strategies, is there still a role for blanket lockdowns? On the one hand, blanket lockdown strategies can reduce burdens on hospital systems, particularly in the initial phase, [50, 59], while on the other hand, the rebound effects may still induce substantial strains on hospital systems later on (Fig. S6). The excessive costs and rebound effects are robust features of blanket lockdowns, both in our model (Fig.S4 & Fig. S7) and confirmed in previous studies, e.g., [60, 61, 11]. The rebound size in our model is also large—nearly 100% of cases averted during the blanket lockdown reoccur later on. While “targeted lockdowns” that lock down areas or businesses burdened with higher transmission rates [62, 63] avoid some of the excess costs of blanket lockdowns, they are still blunt instruments compared to targeted isolation. Nonetheless, blanket lockdowns and targeted isolation strategies may be complementary—reducing hospital strains and managing rebound effects by correcting the coordination problem.

As vaccines are being deployed, new SARS-CoV-2 variants are now circulating in many countries [64]. Thus it continues to be critical to avoid premature relaxation of disease-economy management measures [30]. Our results carry insights for vaccine deployment, to the extent vaccination limits infectiousness. Since our model shows that one infectious individual failing to isolate will induce many susceptible individuals to withdraw, our model insights are consistent with prioritizing vaccines to individuals who, when infectious, are least likely or able to isolate (and therefore most likely to contribute to spread). Using targeted isolation throughout vaccine delivery can further reduce economic costs and disease burden.

There remain many opportunities and open challenges in coupled-systems modelling of disease control and economy management. There is important heterogeneity in transmission, infectiousness, and exposure (e.g., superspreading events and crowding [65, 66]), though explicitly incorporating such heterogeneity into the coupled systems is non-trivial. As greater amounts of high-fidelity mobility data become available, it is important to build data-driven mappings between mobility, contacts, and economic activities within transmission models—[67, 68, 10, 69] offer promising steps in this direction. However, connecting mobility to contact rates and infection probabilities (given a contact) will require further consideration. Finally, it is critically important to consider how to design incentives to implement targeted isolation programs which can sustain participation and compliance.

As an endgame strategy, targeted isolation could avert trillions in recessionary losses while effectively controlling the epidemic. Put differently, disease-economy trade-offs are inevitable when the coordination failure cannot be resolved. Amidst the ongoing public policy debate about economic relief, lockdown fatigue, and epidemic control [26], allocating funds to solving the coordination problem— providing incentives for infectious individuals to learn their disease status and isolate—likely passes the cost-benefit test.

## Materials and Methods

Here we provide an overview of the key elements of our framework including describing the contact function that links economic activities to contacts, the SIRD model, the dynamic economic model governing choices, and calibration. The core of our approach is a dynamic optimization model of individual behavior coupled with an SIRD model of infectious disease spread. Additional details are found in the SI.

## Coupled epi-economic model

### Contact function

We model daily contacts as a function of economic activities (labor supply, measured in hours, and consumption demand, measured in dollars) creating a detailed mapping between contacts and economic activities. For example, all else equal, if a susceptible individual reduces their labor supply from 8 hours to 4 hours, they reduce their daily contacts at work from 7.5 to 3.75. Epidemiological data is central to calibrating this mapping between epidemiology and economic behaviour. Intuitively, the calibration involves calculating the mean number of disease-transmitting contacts occurring at the start of the epidemic and linking it to the number of dollars spent on consumption and hours of labor supplied before the recession begins.

We use a SIRD transmission framework to simulate SARS-CoV-2 transmission. This is supported by several studies (e.g., [70, 71]) that identify infectiousness prior to symptom onset. We consider three health types *m* ∈{*S, I, R*} for individuals, corresponding to epidemiological compartments of susceptible (*S*), infectious (*I*), and recovered (*R*). Individuals of health type *m* engage in various economic activities 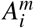, with *i* denoting the activities modelled. One of the 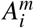 is assumed to represent unavoidable other non-economic activities, such as sleeping and commuting, which occur during the hours of the day not used for economic activities (see SI). Disease dynamics are driven by contacts between susceptible and infectious types, where the number of susceptible-infectious contacts per person is given by the following linear equation:

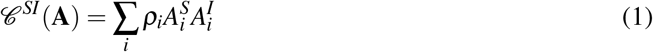

While similar in several respects to prior epi-econ models [15, 16, 67], a methodological contribution is that *ρ*_*i*_ converts hours worked and dollars spent into contacts. For example, *ρ*_*c*_ has units of contacts per squared dollar spent at consumption activities, while *ρ*_*l*_ has units of contacts per squared hour worked.

We also consider robustness to different functional forms in figure 4F & G as a reduced-form way to consider multiple consumption and labor activities with heterogeneous contact rates. Formally:

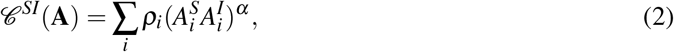

where *α* > 1 (convex) corresponds to a contact function where higher-contact activities are easiest to reduce or individuals with more contacts are easier to isolate. *α* < 1 (concave) corresponds to a contact function where higher-contact activities are hardest to reduce or individuals with fewer contacts are easier to isolate. The baseline case (*α* = 1) implies all consumption or labor activities and individuals have identical contact rates (See SI section 2.3.2 for further discussion and intuition).

### Calibrating contacts

To calibrate the contact function, we use U.S.-specific age and location contact matrices generated in [6], which provide projected age-specific contact rates at different locations in 2017 (shown in SI section 2.3.1). We group these location-specific contact matrices into matrices for contacts during consumption, labor, and unavoidable other activities. The transmission rate was calibrated to give a value of *ℛ*_0_ = 2.6, reflective of estimates [72]. For this, we use the next-generation matrix [36]. The next-generation matrix describes the “next generation” of infections caused by a single infected individual; the *ℛ*_0_ is the dominant eigenvalue of the next-generation matrix (see SI). This calculation is done at the disease-free steady state of the epidemiological dynamical system, when all the population is susceptible. Specifically, we calculate the benchmark number of contacts from each activity in the pre-epidemic equilibrium (e.g., 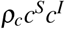 for consumption from equation 1), under pre-epidemic consumption and labor supply levels. We then calculate the coefficients *ρ*_*c*_, *ρ*_*l*_, *ρ*_*o*_ (for consumption, labor, unavoidable other) using 1 such that pre-epidemic consumption and labor supply levels equal the benchmark number of contacts. To account for contacts that are not related to economic activities, the “unavoidable other” contact category is normalized to 1, so that the coefficient *ρ*_*o*_ is simply the number of contacts associated with unavoidable other activities. While pre-pandemic contact structures are necessary to calibrate *ℛ*_0_, our model allows contacts to evolve over time as a function of individual choices, which respond to disease dynamics.

The contact matrices in [6] measure only contacts between individuals in different age groups by activity, without noting which individuals are consuming and which are working. Given the lack of precise data on contacts between individuals engaging in different activities, we simplify by assuming individuals who are consuming only contact others who are consuming, and individuals who are working only contact others who are working. However, in reality individuals who are consuming also interact with individuals who are working (e.g., a bar or restaurant). Future work could collect more detailed contact data describing contacts between individuals engaging in different activities.

### SIRD epidemiological model

The SIRD model is given by:

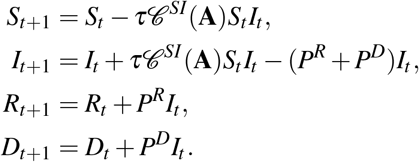

where *S, I, R, D* represent the fractions of the population in those compartments. Because the contact function *𝒞*^*SI*^(**A**) returns the number of contacts per person as a function of activities **A**, then *τ* is a property of the pathogen that determines the infections per contact. This decomposes the classic “*β*” in epidemiological modeling into a biological component that is a function of the pathogen (*τ*) and a behavioral component linked to economic activity (*𝒞* (**A**)), such that *β* = *𝒞* (**A**)*τ* (e.g., [16]).

A key input into individual decision making is the probability of infection for a susceptible individual, which per the SIRD model above depends on the properties of the pathogen, contacts generated through economic activities, and the share of infectious individuals in the population:

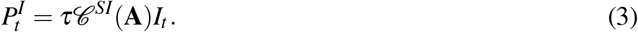

If a susceptible individual reduces their activities (and thus contacts) today, they reduce the probability they will get infected, which in turn reduces the growth of the infection. However, if they keep their economic behavior the same, they enjoy those benefits today, but take the risk of becoming infected in the future. Finally, *P*^*R*^ is the rate at which infectious individuals recover, and *P*^*D*^ is the rate at which they die. Both are assumed to be constant over time and independent of economic activities and contacts.

Our framework can be generalized to other structured compartmental models beyond mean-field (homogeneous) SIRD models. The key feature to translate is the contact function. For example, in an age-structured model the contact function would need to reflect age-specific consumption and labor supply patterns.

### Choices

In order to analyze the three control strategies (voluntary isolation, blanket lockdown, targeted isolation), we solve two types of constrained optimization problems: a decentralized problem and a social planner problem. The decentralized problem reflects atomistic behavior by individuals—they aim to maximize their personal utility and make choices regarding economic activity. The decentralized problem is used to analyze the voluntary isolation and blanket lockdown strategies. Conversely, in the social planner problem, a social planner considers the utility of the population as a whole and coordinates economic activity to jointly maximize the utilities of all individuals in the population. Importantly, the social planner internalizes the full economic costs to the population associated with disease transmission. The social planner problem is used to analyze the targeted lockdown strategy.

In the decentralized problem, individuals observe the disease dynamics, know their own health state, and make consumption and labor choices in each period accounting for the risks incurred by contacts with potentially infectious individuals. Let 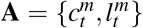 represent the economic activities of consumption and labor chosen in period *t* by individuals of health type *m*. Individuals maximize their lifetime utility by choosing their economic activities, 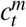 and 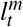, accounting for the effects of infection and recovery on their own welfare:

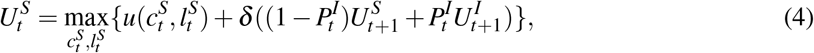

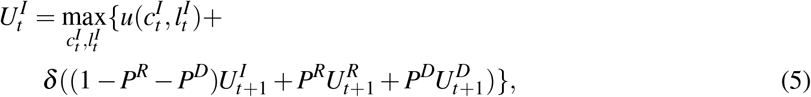

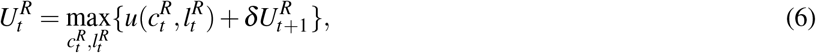

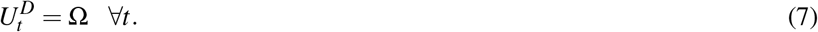

Per-period utility 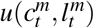 captures the contemporaneous net benefits from consumption and labor choices. In particular, susceptible individuals in period *t* recognize their personal risk of infection 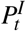 is related to their choices regarding economic activity 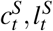, and if they do become infected in period *t* + 1, they have some risk of death in period *t* + 2. Death imposes a constant utility of Ω, calibrated to reflect the value of a statistical life (see SI 2.1.3). The daily discount factor *δ* reflects individuals’ willingness to trade consumption today for consumption tomorrow.

Finally, individuals exchange labor (which they dislike), for consumption (which they do like) such that their budget balances in each period:

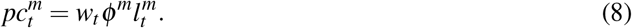

The wage rate *w*_*t*_ is paid to all individuals, per effective unit of labor 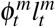, and is calculated from per-capita GDP. The parameter *φ*^*m*^ captures labor productivity such that *φ* ^*S*^ = *φ* ^*R*^ = 1, while *φ* ^*I*^< 1, reflecting the average decrease in productivity of infectious individuals (both asymptomatic and pre-symptomatic [24]—see SI 2.1.1). Following standard practice, the price of consumption *p* is normalized to 1. Finally, market equations that state how individuals are embedded in a broader economy are described in the SI.

The social planner problem coordinates the economic activities of the individuals described above. Instead of economic activities being individually chosen to maximize personal utility, the social planner coordinates consumption and labor choices of each type 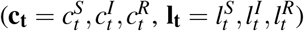 to maximize the utility of the population over the planning horizon, subject to the disease dynamics **??** and budget constraints 8:

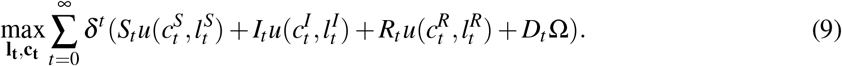

Additional structure (e.g., age compartments, job types, geography) can be incorporated here either by creating additional utility functions or by introducing type-specific constraints. For example, with age compartments, each age type would have a set of utility functions like equations 4-7. These would then be calibrated to reflect age-specific economic activity levels, structural parameters, and observed risk-averting behaviors.

Both the decentralized problem and the social planner problem are solved for optimal daily consumption and labor supply choices in response to daily state variable updates, and we normalize the total initial population size to 1 for computational convenience.

### Utility calibration

Details of the utility function calibration and data sources are found in the SI. Briefly, economic activity levels and structural economic parameters are calibrated to match observed pre-epidemic variables for the US economy. We calibrate risk aversion and the utility cost of death to match the value of a statistical life. This approach ensures both the levels of economic choice variables and their responses to changes in the probability of infection are consistent with observed behaviors in other settings.

## Data Availability

Data and code for reproducibility will be available upon request. Will be available on a GitHub repository upon acceptance

