## Supplementary material for "Disease-economy trade-offs under alternative pandemic control strategies": SI

February 12, 2021

### 1 Model overview

We calculate the economic costs and incidence (infected individuals per 100,000) under different control strategies in three steps. First, we calibrate the utility functions, budget constraints, and contact functions to match pre-epidemic economic and contact data. We choose parameters governing the disease dynamics based on estimates from the literature on COVID-19. All calibrated parameters can be found in Tables S3 and S4. Using the calibrated functions, we solve the model and generate daily time paths of the economic and disease outcomes under voluntary isolation, targeted isolation, and blanket lockdowns for approximately 1.5 years (548 days). Finally, we calculate the economic costs as a discounted sum of consumption losses under each scenario and the cases per 100,000 from the final state of each scenario.

This coupled-systems model has two core components:

1. **Utility functions and budget constraints:** Individuals choose to maximize their utility functions subject to their budget constraints. A key component of individuals' decision-making is that they choose activity levels with the awareness of the probability they get infected at different activities, and respond accordingly. Their choices determine the probability of infection at different activity sites.

2. **Disease dynamics:** The disease evolves as a function of individuals' choices and the prevalence of individuals with different disease statuses. These in turn feed back into individuals' choices.

### 2 Model components and calibration

#### 2.1 Utility functions

The individual per-period utility function $u(c,l)$ in equations 3-5 consists of several pieces. First, the per-period utility from consumption and labor $\hat{u}(c,l)$ has the following functional form:

$$\hat{u}(c,l) = ((\alpha^{\frac{1}{\sigma}} c^{\frac{\sigma-1}{\sigma}} + (1-\alpha)^{\frac{1}{\sigma}} (1-l)^{\frac{\sigma-1}{\sigma}})^{\frac{\sigma}{\sigma-1}})(1+\upsilon). \tag{1}$$

This function represents individual preferences over the economic activities of consumption and leisure/labor and is a standard Constant Elasticity of Substitution (CES) utility function. Individuals value consumption of goods and services $c$ and their leisure (non-labor) time $(1-l)$. This functional form allows us to calibrate how much the individual values consumption relative to leisure ($\alpha$) and the elasticity of substitution between consumption and leisure ($\sigma$). $\upsilon$ measures the utility of unavoidable other contacts, such as those occurring at home. Since unavoidable other contacts do not vary over time and are common to all individuals and health types, we normalize $\upsilon$ to 0 without loss of generality.

Finally, individuals are averse to uncertainty (risk aversion)—they are willing to exchange things they value (e.g., consumption) to reduce the uncertainty associated with becoming infected. Note that this is distinct from their preference for not getting infected; while individuals prefer to remain healthy, risk aversion captures their preference for certainty. In economic terminology, individuals wish to "consumption smooth" across susceptible, infected and other states of the world. This is modeled via the standard Constant Relative Risk Aversion (CRRA) functional form. Putting equation 1 into the CRRA format gives the full per-period utility function as:

$$u(c,l) = \frac{\hat{u}(c,l)^{1-\eta} - 1}{1-\eta} \tag{2}$$

The risk-aversion parameter $\eta$ plays an important role in economic models and governs the degree of risk-averting behavior individuals undertake. If $\eta < 0$, individuals seek out the uncertainty associated with risks; if $\eta = 0$, individuals are indifferent to uncertainties associated with risks; and if $\eta > 0$ individuals try to avoid the uncertainties associated with risks. Following a long literature on risk aversion (e.g., [1]), we take $\eta > 0$ in all model simulations. We discuss this further in section 2.1.2.

We perform a grid search to calibrate utility parameters ($\sigma$ and $\alpha$) and the hourly wage ($w$) so as to reproduce benchmark values of consumption demand ($c^*$ in Table S3), labor supply ($l^*$ in Table S3), and uncompensated wage elasticity of labor supply (0.15, consistent with the compensated elasticities of broad income and income effects in [2]). In calibrating the daily labor supply, we assume time spent sleeping is not counted as leisure. The yearly consumption calibration is annuitized to a daily scale, implying individuals are also making labor-leisure tradeoffs on weekends to facilitate consumption.

We set the daily endowment of time usable in consumption, labor, and leisure to 12 hours. The rest of the day is assumed spent sleeping, commuting, and performing other unavoidable non-economic activities. This implies that in the pre-epidemic equilibrium, 8 hours of the day are spent working and the remaining 4 hours are allocated to consumption and leisure. Unavoidable other contacts are assumed to occur during the remaining 12 hours of the day which are not usable for economic activities.

###### 2.1.1 Productivity and asymptomatic/pre-symptomatic individuals

We do not explicitly model asymptomatic status, pre-symptomatic stages, or mild infections in individuals. We assume infected individuals are unable to work only if they present severe symptoms. Infected individuals who are asymptomatic, pre-symptomatic, or have mild infections, consume and work as though they have no knowledge they are infected.[1] The pathogen-specific productivity parameter $\phi_I$ converts hours worked by average infected individuals into output. We assume only asymptomatic/ pre-symptomatic individuals and those experiencing mild symptoms are able to work, so that $\phi_I$ reflects the share of the infected population which is able to work at any given time. Thus, $\phi_I$ determines the quantity of asymptomatic individuals for a given pathogen.

We calculate $\phi_I$ as a duration-weighted average of time spent in states of differing infection severity. Letting the proportion of the population which is asymptomatic or pre-symptomatic be $P_a$, the proportion of minor sufferers be $P_m$, the proportion of major sufferers be $P_M$, the proportion of infected time spent symptomatic be $t_s$, and the proportion of infected time spent asymptomatic

[1]To emphasize, all individuals in our model are aware of their infection status. It has been shown that a vast majority of transmission events occurs with asymptomatic/pre-symptomatic individuals [3, 4]; with full information there is no distinction between symptomatic and asymptomatic/pre-symptomatic infectious individuals in the compartmental structure of the model.

or pre-symptomatic be $t_a$, this yields

$$\phi_I = 1 * (P_a + P_m) + (0 * t_s + 1 * t_a) * P_M. \tag{3}$$

Assuming major sufferers are hospitalized gives $P_M = H$ (the percentage of those infected that are hospitalized), and the proportion of infected time spent asymptomatic is $t_a = d_a/d_h$; $d_a$ is the duration spent asymptomatic or pre-symptomatic and $d_h$ is the overall duration infected for an individual that is hospitalized. Applying the values from Table S4 gives $P_M = 0.199$, $P_a + P_m = 1 - 0.199$, $t_a = 0.2737$, $t_s = 1 - 0.2737$, giving the average daily productivity while infected as $\phi_I = 0.8555$.

Note that equation 3 can imply negative asymptomatic and minor sufferer shares when $\phi_I$ falls below $t_a$, or shares greater than 1 when $\phi_I$ goes above $1 - t_a$. That is, inverting equation 3 yields

$$P_a + P_m = \frac{\phi_I - t_a}{1 - t_a}. \tag{4}$$

Given our strategy to calibrate the productivity while infected, the physically-plausible range over which $\phi_I$ can vary is therefore $[t_a, 1]$, or $[0.2737, 1]$—implying a productivity *loss* that must be in $[0, 0.7263]$.

Negative shares of asymptomatic/ pre-symptomatic and minor sufferers are possible because the share of time spent asymptomatic or pre-symptomatic ($t_a$) does not vary smoothly with the share of individuals who are likely to transit to that state ($P_a + P_m$). While the share of individuals *currently* in the asymptomatic/pre-symptomatic or minor sufferer states will depend on the share of time spent in those states, $P_a$ and $P_m$ are the probabilities of transiting to those states, not the proportion of individuals in them. To avoid the problem of negative shares, the time spent in asymptomatic/pre-symptomatic or minor sufferer states must go to 0 when the proportion of pre-symptomatic or asymptomatic and minor sufferers are 0 (i.e., $P_a + P_m = 0 \implies t_a = 0$), and vice versa (i.e., $P_a + P_m = 1 \implies t_a = 1$). Given the current lack of knowledge about how the probability an individual is asymptomatic or a minor sufferer relates to the time they are likely to spend asymptomatic, more detailed functional relationships would require speculation. Future work better mapping the structural features of COVID-19 will be necessary to construct such relationships.

##### 2.1.2 Risk aversion

Risk aversion is the extent to which individuals dislike uncertainty when making choices. For example, risk-averse individuals would be willing to accept a lower payoff overall if it meant they were to face no COVID risk whatsoever. The degree to which individuals have this preference for no risk is governed by the parameter $\eta$ (the coefficient of relative risk aversion) in equation 2. The sensitivity of model projections to $\eta$ is explored in section 4.5.

Calibrating $\eta$ for daily-level decisions under risks with potentially severe stakes (e.g., a probability of death) and probabilities that change daily presents many challenges. The most notable is the "calibration theorem" established in [5]: appreciable degrees of risk aversion over moderate stakes implies near-risk neutrality when the stakes are small, and unrealistically high degrees of risk aversion over larger stakes. That is, a moderate coefficient of relative risk aversion (e.g., $\eta$ of 1) does not fit a pandemic setting where extreme negative outcomes (e.g., death) exist and the daily probability of infection may vary over orders of magnitude. Consequently, we do not use a coefficient of relative risk aversion near 1, which is consistent with annual or quarterly decision-making and common in the literature on financial risks [6].

Instead, we use a coefficient of relative risk aversion of 0.1 to reflect low levels of risk aversion at a daily scale. We argue such a low value of risk aversion at a daily scale is plausible for two reasons. First, as shown in [6], the coefficient of relative risk aversion is related to the ratio of the income effect to the price effect. Our low $\eta$ therefore implies that at the daily level, income effects are smaller relative to price effects than they are at a quarterly or annual frequency. Intuitively, this means daily changes in hours worked are more sensitive to wage changes than to changes in unearned income.

Second, research shows that a lower $\eta$ of 0.1 for daily decisions is consistent with higher $\eta$ of 1 for less frequent, less familiar decisions (e.g., financial decisions). [1] finds that less-familiar commitments induce local risk aversion (over moderate-scale income fluctuations) which is an order of magnitude larger than global risk aversion (over large-scale income fluctuations). Infection and death are potentially large-scale income fluctuations. This justifies a lower $\eta$ for daily decisions which could result in infection and death as consistent with a higher value of $\eta$ for less-frequent, lower-stakes decisions around financial risks.

However, while we use a low value of $\eta$ based on the arguments above, the exact value of $\eta$ does not qualitatively change our results. We calibrate the utility of death to obtain a Value of a Statistical Life (VSL) of $10 million USD (consistent with [7] and [8]). As shown in equation 11, for any plausible value of $\eta$ there exists a utility of death which achieves the target VSL. That is, it is $\eta$ **and** the utility of death (the size of the stakes) that jointly determine the resulting risk-averting behavior. Thus for any plausible choice of $\eta$, by setting the utility of death to obtain a desired VSL, we are able to calibrate the model to reproduce observed trade-offs between consumption and risk of death.

##### 2.1.3 Value of a Statistical Life (VSL)

Since the relevant risk of death is faced by infected individuals, we calibrate the utility associated with death so that infected individuals exhibit a VSL of \$10 million, which is consistent with recent estimates from choices to trade risk of death for money in other settings [7, 8]. Since infected individuals are the ones directly facing the risk of death, the utility parameters reflecting VSL for infectious diseases such as COVID ought to be calibrated using the infected individuals' lifetime utility. This then influences the susceptible individuals' choices through the probability of transitioning to the infected state. We approximate the number of life-days remaining for the average individual in each state as infinity. The lifetime utility of being recovered is

$$U^R = \sum_{t=0}^{\infty} \beta^t u(c_R, l_R) \tag{5}$$

$$= \frac{u(c_R, l_R)}{1-\beta}. \tag{6}$$

Let the utility associated with death due to infection be a constant, $\Omega$.[2] The lifetime utility of being infected is as below. It is composed of the utility obtained today, while infected, plus the discounted remaining lifetime payoffs if another state such as recovered or death is entered. Each of these are weighted by their probabilities of occurring in that period, e.g., $P^R, P^D$, etc.

$$U^I = u(c_I, l_I) + \beta \left[P^R U^R + P^D \Omega + (1 - P^R - P^D) U^I\right] \tag{7}$$

$$U^I - \beta(1 - P^R - P^D) U^I = u(c_I, l_I) + \beta \left[P^R U^R + P^D \Omega\right] \tag{8}$$

$$U^I = \frac{u(c_I, l_I) + \beta \left[P^R U^R + P^D \Omega\right]}{1 - \beta(1 - P^R - P^D)} \equiv U^I(c_I, P^D, \Omega). \tag{9}$$

[2] While the utility associated with living states can be measured using choices between streams of consumption in different states, the utility associated with death is more difficult measure. Preferring a particular stream of consumption over death indicates an upper bound, and preferring to die rather than experience a lower stream of consumption indicates a lower bound. Repeated rejections of consumption streams in favor of death can provide more precise estimates but can be hard to observe.

The VSL is a ratio of additional consumption and additional risk of death, $\frac{\Delta c}{\Delta p}$, satisfying the following equation. This follows since the monetary value of life is inferred from how much individuals are willing to pay (measured as a reduction in consumption, $\Delta c$) to avoid an increased chance of death ($\Delta p$).

$$U^I(c+\Delta c, P^D+\Delta p, \Omega) = U^I(c_S, P^D, \Omega). \tag{10}$$

A VSL of \$10 million implies a compensation of \$1,000 for an additional 1/10,000 chance of death. We then set the utility associated with death, $\Omega$, to be consistent with this:

$$\Omega : U^I(c_I+1000, P^D+1/10000, \Omega) = U^I(c_I, P^D, \Omega). \tag{11}$$

The $\Omega$ which satisfies equation 11 will depend on utility parameters, including the coefficient of relative risk aversion. With $\eta = 0.1$ the utility of death implied by a VSL of \$10m is approximately $-672,827$ utils. For any plausible value of $\eta$ (between 0 and 1.25, based on elasticity estimates in [6]), there exists a value of $\Omega$ which satisfies equation 11.

The dependence between $\eta$ and $\Omega$ can be striking. To see the implications, consider the difference between the VSL implied by using $(\Omega, \eta) = (0, 1)$ (a commonly-used combination in this literature, e.g., [9]) and the VSL implied by using $(\Omega, \eta) = (0, 0.1)$. In the former case, the VSL implied by solving equation 10 for $\Delta c$ (given $\Delta p = 1/10000$) is around \$174.5m. In the latter, the implied VSL is around \$2.6m. While neither is near the range of recent VSL estimates, the VSL produced by $(\Omega, \eta) = (0, 1)$ is an order of magnitude larger than the current consensus estimate of \$10m. We explore the implications of this dependence for individual recessionary losses averted and case ratios between the decentralized and planner's management in section 4.5.

#### 2.2 Markets

Individuals in the model are embedded in a broader economy. Similar to [9] and other models of choice in general equilibrium, we assume individuals in this economy supply labor, which is used to produce a good which they consume. Markets clear such that:

$$\begin{aligned} C_t &= \theta L_t \\ \Pi_t &= C_t - w_t L_t = 0 \\ S_t c_t^S + I_t c_t^I + R_t c_t^R &= C_t \\ S_t l_t^S + I_t l_t^I + R_t l_t^R &= L_t \end{aligned} \tag{12}$$

The first two equations in 12 represent standard economic assumptions that firms demand labor $L_t$ to convert into consumption goods $C_t$ at rate $\theta$, and that firms sell those goods and pay for labor until zero economic profits are realized. Since markets are competitive, $w = \theta$ for healthy workers and $w = \phi_I \theta$ for infected workers (i.e., workers are paid their marginal product, and healthy workers are on average more productive than infected workers).

The final two equations in 12 state that total consumption demanded across health types ($\sum_{h \in \{S,I,R\}} h_t c_t^h$) equals total consumption supplied ($C_t$), and that total labor supplied across health types ($\sum_{h \in \{S,I,R\}} h_t l_t^h$) equals total labor demanded ($L_t$).

#### 2.3 Disease dynamics

Disease dynamics follow a homogeneous (mean-field) SIRD compartmental model, where individuals transition from susceptible (S) to infected/infectious (I) states if they get infected by contact with an infected person, and from infected either to recovered (R) or dead (D) states. The parameter values for the SIRD model are drawn from the literature, and reflect mean estimates for COVID-19. The SIRD model is given by:

$$\begin{aligned} S_{t+1} &= S_t - \tau \mathscr{C}^{SI}(\mathbf{A}) S_t I_t, \\ I_{t+1} &= I_t + \tau \mathscr{C}^{SI}(\mathbf{A}) S_t I_t - (P^R + P^D) I_t, \\ R_{t+1} &= R_t + P^R I_t, \\ D_{t+1} &= D_t + P^D I_t. \end{aligned} \tag{13}$$

To calibrate the spread of the disease, we set two types of parameters: the contact rates at activities (one parameter per activity), and the number of infections per contact (a single parameter across all activities). We initialize our model with one infected individual per million. We use parameter estimates of the infection as listed in Table S3, in particular using estimates from [10, 11] and mortality data for the United States from the Centre for Evidence-based Medicine [12].

##### 2.3.1 Contact rates

The contact function converts economic activity levels (e.g., amounts of consumption and labor) between susceptible and infected individuals into daily contacts (number of people encountered in the course of these activities, representing opportunities for infection). This function provides a ready-to-calibrate mapping between economic choices and the resulting outcomes relevant for disease transmission (contacts), and is the key methodological innovation of this paper. Re-writing equation 1 in the main text:

$$\mathscr{C}^{SI}(c,l,o) = \rho_c c_S c_I + \rho_l l_S l_I + \rho_o. \tag{14}$$

The function has three parameters: $\rho_c$ (contacts per squared dollar spent), $\rho_l$ (contacts per squared hour worked), $\rho_o$ (daily unavoidable contacts). For example, suppose an infected individual consumes \$10 in a given day and a susceptible individual consumes \$20. The product of these two provides a measure of how frequently these two groups interact in the course of the consumption activity. The parameter $\rho_c$ converts this into units of contacts. These parameters are common for contacts between any two types, e.g., between susceptibles or between susceptible and infected individuals. We set these parameters to reflect the age-structured daily average contacts from the matrices in [13] as described below.

The matrices in [13] are synthetic contact matrices that estimate age- and location-specific contacts for several countries. We use the matrices generated for the US population in 2017. They cover contacts at work, school, home, and other sites for individuals in 5 year intervals 0-80 then 80 and older. We classify "work" and "school" contacts as occurring during labor activity, "other" contacts as occurring during consumption activity, and "home" contacts as the daily unavoidable residual contacts. We classify "school" contacts as labor since it is a work site for teachers, staff, and other workers.[3] Using synthetic contact matrices allows us to reflect the additional contacts and mixing induced by students as described below. Figure S1 shows the consumption, labor, and unavoidable contact matrices we construct from those given in [13].

In the pre-epidemic equilibrium, the number of contacts between two susceptible individuals (since at this point no infected individuals exist) at consumption sites, labor sites, and unavoidable activities are

[3]It seems reasonable to determine the classification of school sites by the way employees relate to them. Students typically do not attend school when on-site school employees are absent, while school employees do show up at school premises when students are absent.

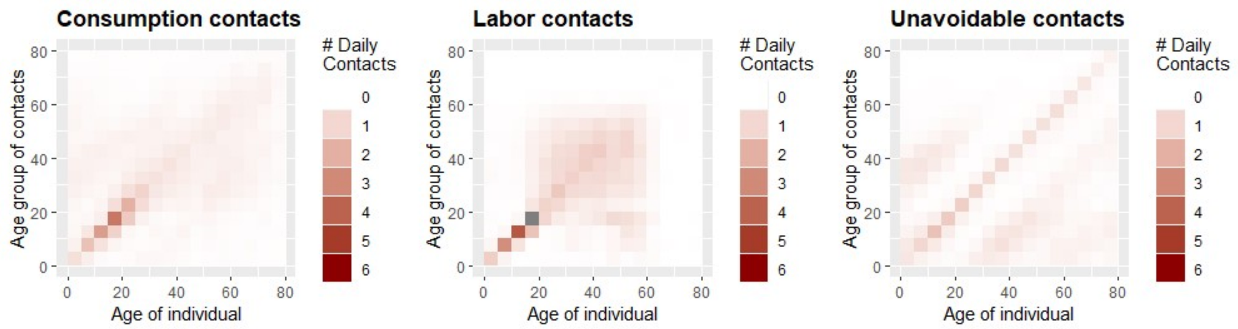

**Figure S1:** Contact matrices: daily contacts by economic activity and by age group. Constructed from [13].

$$\mathscr{C}^{SS}(c) = \rho_c c_S c_S \tag{15}$$
$$\mathscr{C}^{SS}(l) = \rho_l l_S l_S \tag{16}$$
$$\mathscr{C}^{SS}(o) = \rho_o. \tag{17}$$

We estimate pre-epidemic $C^{SS}(a), \forall a \in \{c, l, o\}$ using the matrices from [13] and set pre-epidemic $c_S, l_S$ from our calibration in section 2.1. Using these, we calculate the contact coefficients $\rho_c, \rho_l, \rho_o$ consistent with pre-epidemic economic activity levels and contact rates. Movement of the model's key mechanism, the mapping between economic choices and contacts, is therefore conditioned on relevant and detailed epidemiological data.

We use the next generation matrix method to calculate the mixing-adjusted average number of contacts (e.g., $C^{SS}(c)$) at each type of site as the dominant eigenvalue of the sum of the associated matrices. This approach ensures the contact rates are epidemiologically appropriate for calibrating $\mathscr{R}_0$. In section 2.3.3 we provide a brief primer on this approach—we refer readers to [14] for a more thorough discussion of the method.

##### 2.3.2 The shape of the contact function

Our main results assume a standard linear contact function, shown in equation 14, which implies all activities and individuals have the same rate of contacts per unit of activity. However, individual heterogeneity in contact patterns by activity can affect both disease transmission and economic outcomes. For example, superspreading events occur when specific activities or individuals lead to much higher degrees of transmission than others. As described in the Materials and Methods section of the main text, we vary the shape of the contact function in order to model such heterogeneity. The shapes we consider are described by equation 18.

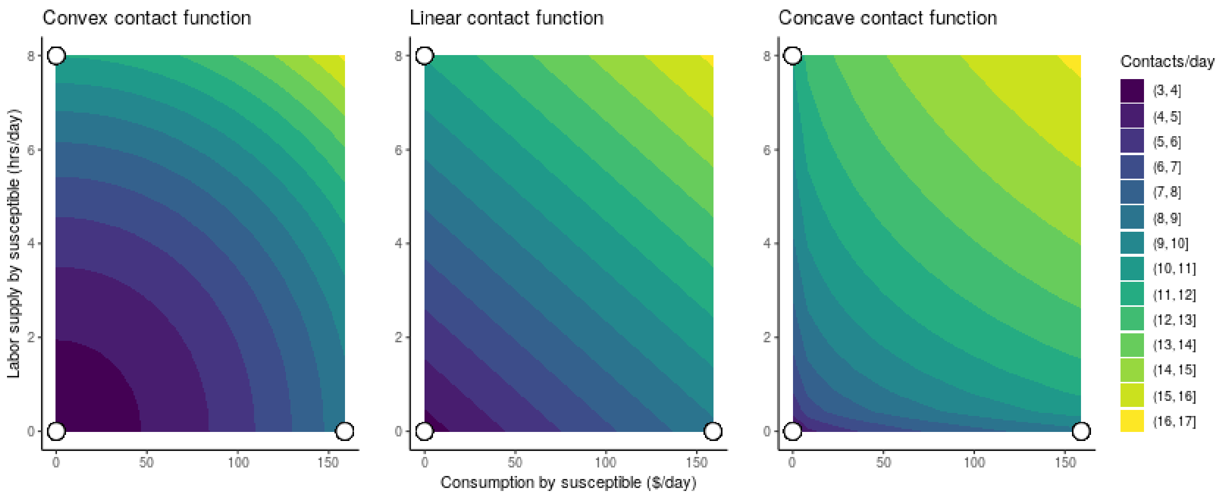

**Figure S2:** Calibrated contact surfaces showing the mapping between economic activity levels and contacts per day. The three white points show where contact matrix data (shown in Figure S1) was used to calibrate the surfaces. Consumption data was used to calibrate the point in the lower right, labor supply data the point in the upper left, and data at unavoidable activities used to calibrate the point at the origin.

$$\mathscr{C}^{SI}(\mathbf{A}) = \rho_c(c^S c^I)^\alpha + \rho_l(l^S l^I)^\alpha + \rho_o. \tag{18}$$

Equation 14 is the case when $\alpha = 1$. The contact function is convex when $\alpha > 1$, and concave when $\alpha < 1$. Either convex or concave forms can represent heterogeneity across activities or individuals, where a few activities or individuals account for the majority of contacts (e.g., a power law distribution of individual contacts). The degree of heterogeneity is captured in how far $\alpha$ is from 1. The key distinction between the convex and concave forms is how the individuals or activities are prioritized for isolation/reduction. A concave form implies high-contact activities or individuals are the last ones to be reduced or isolated, while a convex form implies high-contact activities or individuals are the first ones to be reduced or isolated.

If the costs of isolating individuals or reducing activities were homogeneous, then a social planner would likely prioritize isolation or reduction, inducing a convex contact function. As shown in the main text, such prioritization would increase the gains from targeted isolation. Intuitively, this result arises because with a convex contact function any given reduction in contact levels can be achieved with lower levels of isolation or activity reduction than under a linear contact function.

Calibrating the exponent $\alpha$ in equation 18 is challenging. As shown in figure S2, the pre-pandemic data points we observe can be fit equally well by many surfaces. Additional data on contacts at lower levels of activity prior to the pandemic is required to calibrate $\alpha$. Such data could be obtained by incorporating questions about consumption and labor supply into contact surveys. More importantly, however, the calibration of all contact function parameters will likely need to change over time in order to reflect pandemic-induced changes in the structure of the economy.

Structural changes in the economy in response to the pandemic—such as increasing use of contactless delivery and drop-off, staggered time use of shared office spaces, and the availability of "quarantine hotels"—are likely to shift, tilt, or deform the contact function. To the extent that the pre-pandemic economy features uniformly more contacts, the use of pre-pandemic data (regardless of how many points are used) is likely to overstate the number of contacts associated with higher levels of activity. This is an instance of the Lucas Critique [15]—data prior to a structural change will conflate the impact of policies addressing the change with the effects of the change itself. While we study the sensitivity of our conclusions to such variations in Figure 4 of the main text, we note that correcting for such bias requires either ongoing contact survey data collection throughout the pandemic or a theoretically-principled way of allowing the contact parameters $\rho_c, \rho_l, \rho_o, \alpha$ to vary over time.

##### 2.3.3 The next-generation matrix method

The term "next-generation" comes from the use of "generations" to describe waves of secondary infections which flow from each prior infection. Thus if $\mathscr{R}_t$ describes the reproduction number of the $t$th generation, then $\mathscr{R}_0$ is simply the number of infections generated by the first case (i.e., generation zero) in an entirely-susceptible population. Defining $\mathscr{R}_0$ in a structured compartmental epidemic model involves calculating the expected number of new infections across all infection types. Though we only use a single type of infected individual in our SIR(D) model, this method allows us to account for individuals with different ages and at different activities.

Assume we have a system with multiple discrete types of infected individuals - e.g., male, female, age classes, high risk low risk. We define the *next-generation matrix* as the square matrix $\mathbf{G}$ in which the $i, j$th element of $\mathbf{G}$, $g_{ij}$ is the expected number of secondary infections of type $i$ caused by a single infected individual of type $j$ assuming the population of type $i$ is entirely susceptible. That is, each element of $\mathbf{G}$ is a reproduction number which accounts for who infected whom.

The next-generation matrix has a number of desirable mathematical properties. In particular, since it is a non-negative square matrix, it is guaranteed to have a unique eigenvalue which is positive, real, and strictly larger than all others. This dominant eigenvalue (or spectral radius) is the basic reproduction number, $\mathscr{R}_0$.

To illustrate, consider a simplified directly transmitted disease in a completely susceptible population with two groups, young and old. Define $y$ as the expected number of infected young individuals and $o$ as the expected number of infected old individuals, given contact with a single infected member of the other group. The next-generation matrix is then

$$\begin{bmatrix} y & 0 \\ 0 & o \end{bmatrix}$$

and $\mathscr{R}_0$ is $\sqrt{yo}$. It is worth noting that this is also the geometric mean of the expected number of young and old secondary cases.

##### 2.3.4 The reproductive number

To set the initial reproduction number $\mathscr{R}_0$, we calculate the number of infections per contact, $\tau$, such that

$$\mathscr{R}_0 = \tau \mathscr{C}^{SI}(c,l,o)d, \tag{19}$$

where $d$ is the mean serial interval and $\mathscr{R}_0$ is the target reproductive number. We initialize our model with the initial prevalence, $\varepsilon$, at one in a million. The transmission rate $\tau$ is tuned using the next-generation matrix implied by the calibrated contact function (equation 14) to give a target value of $\mathscr{R}_0$ of 2.6, consistent with estimates [16, 17].

#### 2.4 Solution concepts

We apply two solution concepts: an "equilibrium" and an "optimal plan". As shown in Materials and Methods, the constrained maximization problems which susceptible, infected, and recovered individuals solve in each period in the decentralized cases (voluntary isolation and blanket lockdown) are

$$U_t^S = \max_{c_t^S, l_t^S} \{u(c_t^S, l_t^S) + \delta((1 - P_t^I)U_{t+1}^S + P_t^I U_{t+1}^I)\}, \tag{20}$$
$$\text{s.t. } pc_t^S \leq w^S l_t^S$$

$$\begin{aligned} U_t^I = \max_{c_t^I, l_t^I} \{ & u(c_t^I, l_t^I) + \\ & \delta((1 - P^R - P^D)U_{t+1}^I + P^R U_{t+1}^R + P^D U_{t+1}^D)\}, \end{aligned} \tag{21}$$
$$\text{s.t. } pc_t^I \leq w^I l_t^I$$

$$U_t^R = \max_{c_t^R, l_t^R} \{u(c_t^R, l_t^R) + \delta U_{t+1}^R\}, \tag{22}$$
$$\text{s.t. } pc_t^R \leq w^R l_t^R$$

The utility of dead individuals is simply

$$U_t^D = \Omega \quad \forall t. \tag{23}$$

The planner's problem is

$$\max_{\mathbf{l_t}, \mathbf{c_t}} \sum_{t=0}^{\infty} \delta^t (S_t u(c_t^S, l_t^S) + I_t u(c_t^I, l_t^I) + R_t u(c_t^R, l_t^R) + D_t \Omega) \tag{24}$$
$$\text{s.t., } \forall t, pc_t^S \leq w^S l_t^S \tag{25}$$
$$pc_t^I \leq w^I l_t^I \tag{26}$$
$$pc_t^R \leq w^R l_t^R. \tag{27}$$

All of the maximization problems are also subject to the disease dynamics (system of equations 13).

An equilibrium is a vector of choices for each type, $(c^S, l^S)$,$(c^I, l^I)$,$(c^R, l^R)$, such that individual utilities are maximized subject to budget constraints and disease dynamics, and markets clear (i.e., a joint solution to the systems in 20, 21, 22, 23, subject to system 13 and total consumption equalling total production: $C_t = \theta L_t$, as shown in system of equations 12). This equilibrium is the voluntary isolation strategy. When we study the blanket lockdown scenario, equilibrium behavior is used to generate disease and economic dynamics while the lockdown doesn't bind. Section 4.1 describes how the blanket lockdown scenario is constructed in more detail.

An optimal plan is a vector of choices for each type, $(c^S, l^S, c^I, l^I, c^R, l^R)$, such that aggregate population utility is maximized subject to budget constraints and disease dynamics, and markets clear (i.e., a solution to the system in 24, subject to system 13 and total consumption equalling total production). This optimal plan is the targeted isolation scenario.

The coordination failure is precisely the difference between solving the decentralized problems (which yield the equilibrium) and solving the social planner's problem (which yield the optimal plan). The equilibrium involves each individual type choosing their labor supply and consumption demand having accounted for their personal benefits and costs from those choices, and ignoring how their choices impact others. The optimal plan instead has the planner coordinating individuals' choices to maximize their collective utility, i.e. accounting for how behavior by one impacts benefits and costs received by others.

There is no coordination failure (i.e., no difference between behavior in the equilibrium vs the optimal plan) for recovered individuals because they neither contribute to disease spread nor are impacted by contact with infectious individuals. Infectious and susceptible individuals are where the differences arise. Infectious individuals' equilibrium behavior ignores how their presence in common spaces impacts susceptible individuals, who withdraw activity in response to infection risk. Since susceptible individuals are the majority of the population in a novel epidemic, the optimal plan prioritizes activity by susceptibles and has infectious individuals withdraw activity instead, thus maximizing the population's aggregate welfare. Social welfare functions which do not aggregate individual utilities may produce different results; we discuss this briefly in section 5.4.4, but exploring this point is beyond the scope of this analysis.

#### 2.5 Individual heterogeneity and coordination failures

For analytical tractability and to highlight the effects of the coordination failure, we study a mean-field SIRD model. However, adding individual heterogeneity, e.g., multiple age classes, does not change the fundamental coordination failure or the use of targeted isolation to address it. We illustrate this point below, using a static discrete-choice game theoretic framework for ease of exposition. We focus on Nash equilibria, which are profiles of choices such that no individual can gain by unilaterally changing their choice. We assume throughout that side payments (i.e., transfers between individuals) are possible.

To start, consider a simple stylized game between representative susceptible and infected individuals deciding whether to engage in contactful economic activities (activity level $H$) or remain home (activity level $L$). The susceptible individual gains utility from engaging in activities when the infected individual is not present in the common space (so that there is no risk of infection), while the infected individual gains utility from engaging in activities regardless of what the susceptible individual does. The infected individual's sickness prevents them from enjoying the activity as fully as if they were not infected, so letting the susceptible individual's payoff from activity without risk of infection be 1, the infected individual's payoff is $0 < \phi < 1$. $\phi$ can also be interpreted as productivity loss reflecting the share of asymptomatic individuals as in equation 3. When the individuals do not engage in activity, they get payoffs of 0, and when the susceptible individual risks infection they are worse off than if they had not engaged in activity at all. Letting the susceptible individual's payoff from risking infection to engage in activity be $-1$ and the populations be of equal size, the game is described by the bimatrix in Table S1.

**Table S1:** Mean-field two-player SI activity game

| | | I | |
|---|---|---|---|
| | | L | H |
| S | L | 0,0 | 0,$\phi$ |
| | H | 1,0 | -1,$\phi$ |

Denoting the susceptible individual's activity level by $a^S$ and the infected individual's activity level by $a^I$, the Nash equilibrium is $(a^S, a^I) = (L, H)$. As long as $\phi < 1$ the Pareto optimum is $(H, L)$. Intuitively, it is always in the infected individual's interest to engage in activity so they always choose $H$, forcing the susceptible individual to remain home and choose $L$. However, they would be collectively better off if they could coordinate on having the susceptible individual engage in activity while the infected individual remains home. That is, the Pareto optimum is targeted isolation. This remains true even when we allow for different population sizes and disease dynamics, as long as there are more susceptible individuals than infected individuals (e.g., in the initial phase of a novel epidemic).

Next, expand the game by segmenting the population into two age classes, young ($Y$) and old ($O$). There are four representative individuals, the young susceptible ($S^Y$), the old susceptible ($S^O$), the young infected ($I^Y$), and the old infected ($I^O$). As before, the infected individuals' payoffs $(\phi^Y, \phi^O)$ from engaging in activity are strictly less than the susceptible individuals'. To mimic one feature of COVID-19, suppose $0 < \phi^O < \phi^Y < 1$, so that the old infected individual receives a lower payoff than the young infected individual. The susceptible individuals get a payoff of 1 from engaging in activity as long as no infected individuals are around, and $-1$ from engaging in activity with any infected individuals around. Letting the row player be $S^O$ and the column player be extended to include the three remaining players (with each combination of strategies being reflected as a unique strategy for the composite column player), the game is as described in Table S2.

**Table S2:** Age-structured four-player SI activity game

| | | $I^Y, S^O, I^O$ | | | | | | | |
|---|---|---|---|---|---|---|---|---|---|
| | | L, L, L | H, L, L | L, H, L | L, L, H | L, H, H | H, H, L | H, L, H | H, H, H |
| $S^Y$ | L | 0,0,0,0 | 0,$\phi^Y$,0,0 | 0,0,1,0 | 0,0,0,$\phi^O$ | 0,0,-1,$\phi^O$ | 0,$\phi^Y$,-1,0 | 0,$\phi^Y$,0,$\phi^O$ | 0,$\phi^Y$,-1,$\phi^O$ |
| | H | 1,0,0,0 | -1,$\phi^Y$,0,0 | 1,0,1,0 | -1,0,0,$\phi^O$ | -1,0,-1,$\phi^O$ | -1,$\phi^Y$,-1,0 | -1,$\phi^Y$,0,$\phi^O$ | -1,$\phi^Y$,-1,$\phi^O$ |

Inspection reveals that $(a^{SY}, a^{IY}, a^{SO}, a^{IO}) = (L, H, L, H)$—the susceptible individuals remain home while the infected individuals engage in activity—is a Nash equilibrium. However, the Pareto optimum is $(a^{SY}, a^{IY}, a^{SO}, a^{IO}) = (H, L, H, L)$—the infected individuals remain home while the susceptible individuals engage in activity. The intuition is the same as the two-individual case: infected individuals are better off engaging in activity whether susceptible individuals are present or not, while susceptible individuals are worse off from engaging in activity near infected individuals. Similar results can be derived with $n$ categories of susceptible and infected individuals.

While disease dynamics, varying population sizes, and additional activity choices may lead to different magnitudes of voluntary and targeted isolation for different groups, additional compartmental structure does not change the fundamental nature of the coordination failure. Infected individuals, particularly those who are not suffering from severe symptoms, face incentives to engage in activities. Susceptible individuals have incentives to avoid infected individuals in common spaces. In equilibrium, too many susceptible individuals will remain home, and too many infected individuals will engage in activity. Targeted isolation solves the coordination failure by keeping infected individuals at home while allowing susceptible individuals to engage in activity.

#### 2.6 Parameter values

Tables S3 and S4 list the calibration targets and calibrated parameter values. Table S3 focuses on the targeted equilibrium economic choices, the contact rates, and disease durations. Table S4 focuses on disease and health system parameters.

**Table S3:** Pre-epidemic equilibrium economic choices, implied utility parameters, and contact rates

| Parameter | Description | Source |
|---|---|---|
| **Time endowment, pre-epidemic economic choices, & calibration targets** | | |
| $\bar{l} = 12$ | Daily hours available for labor and leisure | Consistent with [18] and [19]. |
| $l^* = 0.3333$ | % day worked | Consistent with [18] and [19]. |
| $c^* = 58{,}000/365$ | Daily consumption $ | US 2016 GDP per capita / days in year |
| $\varepsilon_l = 0.15$ | Uncompensated elasticity of labor supply | |
| **Calibrated utility and budget parameters** | | |
| $\beta = 0.96^{1/365}$ | Discount factor (daily) | Implies long-run interest rate of 0.04, as in Goulder et al. (2019). |
| $\sigma = 1.6417$ | Elasticity of substitution between consumption and leisure | Calibration targeting $l^*$ and $c^*$; calculations described in section 2.1 |
| $\alpha = 0.2266$ | Utility share of consumption relative to leisure | |
| $\eta = 0.1$ | Baseline coefficient of relative risk aversion | Described in section 2.1.2. |
| $\Omega = -672827$ | Utility of death | Described in section 2.1.3. |
| $w = 19.865$ | Wage for uninfected individual | Implied by $l^*$ and $c^*$. |
| $\phi_I = 0.8555$ | Productivity of average infected individual *(as share of non-infected productivity)* | Calculations described in section 2.1.1. |
| $\phi_S, \phi_R = 1$ | Productivity of average uninfected individuals | No disease impact on susceptibles and recovereds. |
| **Contact parameters** | | |
| $\mathscr{C}(c) = 5.1664$ | Average daily contacts at consumption activities | Calculations described in section 2.3.1. |
| $\mathscr{C}(l) = 7.5131$ | Average daily contacts at labor activities | |
| $\mathscr{C}(o) = 3.5485$ | Average daily unavoidable contacts | |
| $\rho_c = 0.0002$ | Consumption contact conversion factor | |
| $\rho_l = 0.1174$ | Labor contact conversion factor | |
| $\rho_o = 3.5485$ | Unavoidable contact conversion factor | |

**Table S4:** Disease and health system parameters

| Parameter | Description | Source |
|---|---|---|
| **Durations** | | |
| $d = 5.1$ | Duration of infectiousness (days) | Estimate of mean serial interval from [10] |
| $d_a = 5.2$ | Duration asymptomatic (days) | Estimate of mean incubation period from [10]. |
| $d_h = 19$ | Total duration if hospitalized (days) | $d_h = d_a$ + time from symptom onset to hospitalization (3.8 days from Table 2 of Zhang et al. (2020)) + time until hospital discharge (10 days from [20]) = 19 days. |
| **Probabilities** | | |
| $P^{D*} = 0.015$ | Probability of dying conditional on being infected | Estimate of the Infection Fatality Rate provided by the Centre for Evidence-Based Medicine [11].[4] Based on meta-analyses of the best estimates of case numbers and fatalities. Range given of [5.73,5.81]; we take the mean estimate. |
| $P^{R*} = 0.9423$ | Probability of recovering from infection | $1 - P^{R*}$ |
| $P^{D} = 0.0029$ | Probability infected individual dies on a given day | =Prob(leaving I state in given day)AND Prob(dying from infection) = $(1/d)P^{D*}$ |
| $P^{R} = 0.1931$ | Probability of recovering on a given day | =Prob(leaving I state on given day)AND Prob(recovering from infection) = $(1/d)P^{R*}$ |
| $1 - P^R - P^D = 0.8039$ | Probability of remaining in infected state on a given day | $1 - P^R - P^D = 1 - 1/d$. |
| **Other** | | |
| $\varepsilon = 1 * e^{-6}$ | Starting population infected (%) | Initial condition. Implies US infection spread began with around 300 people. |
| $\tau = 0.0314$ | Infection per contact | Calculation described in section 2.3.4. |
| $H = 19.9\%$ | % hospitalized | 13.8% severe disease + 6.1% critical [21]. |

#### 2.7 Empirical validation to observed US economic outcomes during COVID-19

Our model predicts a 66% peak-to-trough decline in GDP under voluntary isolation (shown in Figure 2 of the main text). While our model reports the daily evolution of the economy, most economic data is reported at a quarterly frequency. To compare our results with empirically observed economic data for the US during COVID-19, we aggregate the daily data and present the implied quarterly downturns from our model. We assume the starting level of COVID-19 prevalence in our model (1 in a million) was roughly the level of COVID-19 prevalence in the US around February 20th, 2020.[5] The comparisons of model-predicted and observed peak-to-trough values are shown in the table below. All actual US economic data is in real terms and taken from the Bureau of Economic Analysis' quarterly "Real Gross Domestic Product and Related Measures" tables.

GDP in our model is closest to "Consumption" in the US economic accounts, since we do not model Investment, Government Spending or Net Exports, which are added to Consumption to form GDP. We also show consumption of services, since we do not explicitly model durable goods purchases. Still, in all cases our model produces quarterly economic outcomes that are broadly consistent with what was observed, despite the simplifying assumptions we make. This suggests our model captures the key features of the epidemic and associated economic downturn.

**Table S5:** Model versus actual economic outcomes: peak-to-trough (quarterly frequency)

| | |
|---|---|
| **Actual US economy (BEA)** | |
| GDP | -35% |
| Consumption | -38% |
| Consumption: services | -48% |
| **Model economic outcomes** | |
| Voluntary isolation (Quarterly) | -33% |

We plot our model's economic outputs at a quarterly frequency alongside observed US economic data in Figure S3. Again, our model provides a good description of the economic reality observed with COVID-19. The economic contraction in our model falls slightly faster than actual data, reflecting the difficulty in knowing precisely when infection prevalence in the real world reached the model's initial condition, as well as the fact that we are comparing with voluntary

[5] Varying the starting date changes the comparison, but does not change the qualitative patterns.

isolation rather than a blanket lockdown scenario (since blanket lockdowns were sporadically in place in parts of the US).

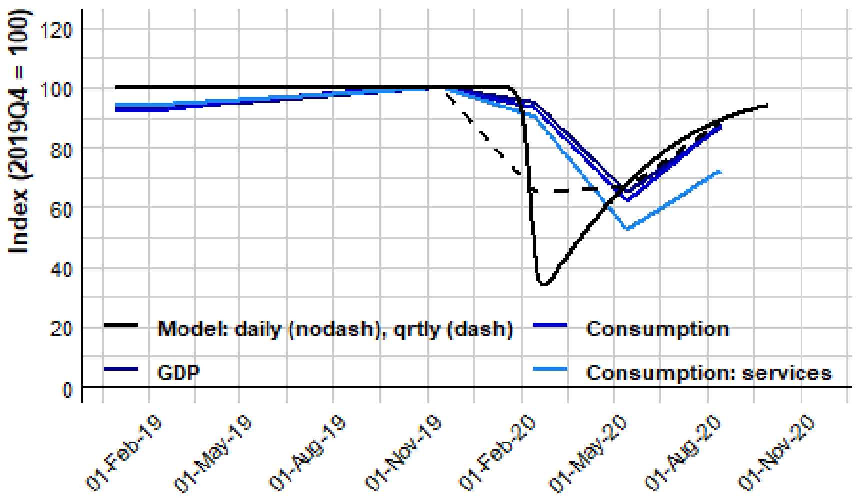

**Figure S3:** Actual economic outcomes under COVID-19 (blue lines) with model equivalents (black lines). Since our model is at a daily frequency, and actual economic data is not produced at daily frequency, we show daily economic outcomes from our model (straight black line) and quarterly economic outcomes (dashed black line).

### 3 Solving the model

In this section we describe some details of how the model is solved. These include the algorithms used to calculate the equilibrium and optimal plan, the construction of the grid on which the model is solved, and a correction to the first-order conditions of the susceptible individual's maximization problem to ensure their choices reflect atomistic behavior.

#### 3.1 Solution methods and algorithms

Decisions about how much to consume and work today depend on the current state of the disease and affect the evolution of disease outcomes. We solve the individual's maximization problem using a dynamic programming algorithm.

The algorithm for finding equilibrium can be broken into three steps. First, we construct an initial guess of the terminal value functions $(U_0^S, U_0^I, U_0^R)$ and use the initial state $(S_0, I_0, R_0)$ with the value function guess to determine the optimal $c, l$ for each type. Within the same time period, we iterate over choices to ensure that all individuals are best-responding to each other—we refer to this as the "equilibrium inner loop". Second, these economic variables are used to determine the contact rate between susceptible and infected individuals according to $\mathscr{C}^{SI}(\mathbf{A}) = \sum_i \rho_i A_i^S A_i^I$. Third, the contacts are plugged into the disease dynamics equations (system 13) to find the next-period state, $\{S_1, I_1, R_1\}$. This procedure is repeated until the value function converges. A more formal description is shown in Algorithm 1.

---

**Algorithm 1:** Algorithm to calculate equilibrium

---

Set

$$U_0(S,I,R) = guess(S,I,R), \tag{28}$$

$$c_0 = c^*, l_0 = l^*, \tag{29}$$

for each type.

Set $i = 1$ and $\delta = 100$ *(some large initial value).*

**while** $\delta > \varepsilon$ **do**

  Set $j = 1$ and $\delta_c = 100$ *(some large initial value)*

  **while** $\delta_c > \varepsilon_c$ **do**

    **for** $h \in \{S, I, R\}$ **do**

      At each grid point in $(grid_S, grid_I, grid_R)$, use a numerical routine to choose $(c_{ij}^h, l_{ij}^h)$ to maximize $U_i^h(S,I,R)$ subject to their budget constraint (as shown in systems 20, 21, 22) and disease dynamics (system 13). When $i = 1$, $U_0^h(S', I', R')$ are used as continuation values; when $i > 1$, $U_{i-1}^h(S', I', R')$ are used. Linear interpolation is used to compute $U_{i-1}^h(S', I', R')$ between grid points.

      $\delta_c \leftarrow ||\sum_h (l_{ij}^h(S,I,R) - l_{ij-1}^h(S,I,R))||$

      $j \leftarrow j + 1$

    **end**

  **end**

  $\delta \leftarrow ||U_i(S,I,R) - U_{i-1}(S,I,R)||_\infty$.

  i ← i+1

**end**

---

The algorithm for finding the optimal plan is simpler, as no equilibrium inner loop is required. It can be broken into similar three steps: First we construct initial value function guesses for each type as in the equilibrium algorithm, then aggregate up to the social welfare function. Using the social welfare function, we then jointly solve for the optimal (social welfare-maximizing) consumption demands and labor supplies for each type. Next we use the choices to determine contacts, and finally use contacts to calculate the next-period state. The procedure is repeated until the social welfare function converges. A more formal description is shown in Algorithm 2.

---

**Algorithm 2:** Algorithm to calculate optimal plan

---

Set

$$U_0(S,I,R) = guess(S,I,R), \tag{30}$$

$$c_0 = c^*, l_0 = l^*, \tag{31}$$

for each type. The social welfare function is then

$$W_0(S,I,R) = S_0U_0^S + I_0U_0^I + R_0U_0^R. \tag{32}$$

Set $i = 1$ and $\delta = 100$ *(some large initial value).*

**while** $\delta > \varepsilon$ **do**

  **for** $h \in \{S,I,R\}$ **do**

    At each grid point in $(grid_S, grid_I, grid_R)$, use a numerical routine to choose $(c_i, l_i)$ to maximize $W_i(S,I,R)$ subject to budget constraints (as shown in system 24) and disease dynamics (system 13). $W_i(S,I,R)$ is given by

$$W_i(S,I,R) = \sum_{h\in\{S,I,R\}} hu(c^h, l^h) + \delta W_{i-1}(S',I',R')$$

    Linear interpolation is used to compute $W_{i-1}(S',I',R')$ between grid points.

  **end**

  $\delta \leftarrow ||W_i(S,I,R) - W_{i-1}(S,I,R)||_\infty$.

  i $\leftarrow$ i+1

**end**

---

We set $\varepsilon$ to 0.05% of the level of the susceptible type's value function for the decentralized problem (algorithm 1), and to 0.001% of the initial social welfare function for the social planner problem (algorithm 2). The equilibrium inner loop occurs in steps 5-11 of algorithm 1 and typically converges within 1-2 iterations. We use the post-epidemic steady-state lifetime utility for each type as the initial value function guesses for each type in both algorithms.

###### 3.1.1 Grid construction

The appropriate choice of grid points is particularly important in this model, since much of the curvature in the susceptible type's choices occur at low levels of infection prevalence. The grid design balances two considerations: an excessively coarse grid provides low fidelity in the initial stages of the epidemic when $S \approx 1$ and $I, R \approx 0$ (resulting in an epidemic which ends very quickly and a smaller recession), but an excessively fine grid incurs substantial computational overhead due to the curse of dimensionality.

We mitigate this tradeoff by using an expanded Chebyshev grid, which has many attractive properties for economic optimization problems [22]. The key property for our purposes is that it provides greater fidelity near the boundaries of the grid (where it is needed) in exchange for lower fidelity in the interior of the grid (where it is less necessary). The formula for the $k^{th}$ expanded Chebyshev node on an interval $[a,b]$ with $n$ points is

$$x_k = \frac{1}{2}(a+b) + \frac{1}{2}(b-a)\sec\left(\frac{\pi}{2n}\right)\cos\left(\frac{k}{n} - \frac{1}{2n}\right).$$

The only physically-meaningful grid points are within the unit 3-simplex $\{(S,I,R,D) : S+I+R \leq 1, \text{ where } S,I,R,D \geq 0, D \leq 1\}$, or equivalently on the face of the unit 4-simplex, $\{(S,I,R,D) : S+I+R+D = 1, \text{ where } S,I,R,D \geq 0\}$. Typical initial conditions for novel epidemics imply an even smaller subset of points is likely to be visited. For simplicity we solve the model on the entire unit hypercube, $\mathbb{R}^4_{[0,1]}$, though more complex methods can exploit the geometry of the problem for more efficient computation. Methods which avoid off-simplex computations will be increasingly valuable as the model is extended to additional dimensions, e.g. including multiple types within each health status.

#### 3.2 Correctly representing atomistic individual behavior

Susceptible individuals solve the following optimization problem:

$$U^S(S,I,R) = \max_{c,l}\{u(c,l) + \beta[P^I(c,l)U^I + (1-P^I(c,l))U^S(S',I',R')]\} \tag{33}$$

$$\text{s.t.} \quad pc \leq wl \tag{34}$$

$$S' = S(1-P^I(c,l)) \tag{35}$$

$$I' = I(1-P^R-P^D) + P^I(c,l)S \tag{36}$$

$$R' = R + P^R I, \tag{37}$$

where we ignore the dead state for brevity since it does not affect the solution, and

$$P^I(c,l) = \tau(\rho_c c c_I + \rho_l l l_I + \rho_o)I$$

is the probability of getting infected interacting with $I$-type individuals at consumption, labor, and unavoidable activities.

The representative individual solving the problem above only faces an externality from $I$ types through $P^I$. There is no externality from the choices of $S$ types since the representative individual controls them all and internalizes this knowledge. This can be seen formally in the first-order condition to the above problem:

$$c^*: 0 = u_c + \beta P_c^I(U^I - U^S) + \beta(1-P_c^I)\frac{\partial U^S}{\partial c} - \lambda \tag{38}$$

$$\implies u_c = \beta P_c^I(U^S - U^I) + \beta(1-P_c^I)SP_c^I(U_S^S - U_I^S) + \lambda \tag{39}$$

and

$$l^*: \lambda = w^{-1}\left(-u_l + \beta P_l^I(U^S - U^I) + \beta(1-P_l^I)SP_l^I(U_S^S - U_I^S)\right) \tag{40}$$

$$\implies (c^*, l^*): u_c = -\frac{u_l}{w} + \beta(P_c^I + \frac{P_l^I}{w})(U^S - U^I) + \beta S\left(P_c^I(1-P_c^I) + w^{-1}P_l^I(1-P_l^I)\right)(U_S^S - U_I^S), \tag{41}$$

where $\lambda$ is the Lagrange multiplier on the budget constraint and subscripts on $P^I$ and $U^S$ denote partial derivatives. However, the final term in equation 41,

$$\beta S\left(P_c^I(1-P_c^I) + w^{-1}P_l^I(1-P_l^I)\right)(U_S^S - U_I^S),$$

contains value function derivatives $U_S^S$ and $U_I^S$ (along with variance terms $P_c^I(1-P_c^I) + P_l^I(1-P_l^I)$). These value function derivatives reflect the representative individual's awareness that changes in the population sizes of susceptible and infected individuals affect the marginal value of remaining susceptible. These terms imply susceptible individuals internalize how their choices might affect each other, which is incompatible with atomistic behavior.

To ensure consistency with atomistic behavior, we instead assume the representative susceptible individual maximizes their value *ignoring* the effect of population size changes on the marginal value of remaining susceptible. This yields the following first-order condition instead:

$$(c^*, l^*): u_c = -\frac{u_l}{w} + \beta(P_c^I + \frac{P_l^I}{w})(U^S - U^I). \tag{42}$$

Equation 42 treats the representative susceptible individual as ignoring how population size changes (which they consider beyond their control) affect the marginal value of remaining susceptible. The above is the activity-structured analog of equation (7) in [23], which does not feature consumption and labor as specific activity classes.

### 4 Sensitivity to parameter variations

In this section we consider model sensitivity to a host of parameter variations.

#### 4.1 Sensitivity to blanket lockdown implementation

To study how blanket lockdowns might compare to the voluntary and targeted isolation policies we focus on, we conduct a sensitivity analysis over plausible lockdowns, informed by US state-level data on lockdowns. We define lockdowns as restrictions on businesses or stay-at-home orders, and define the start of a lockdown as the first date such restrictions are put in place. In all US states, restrictions on businesses weakly preceded stay-at-home orders. We classify the lockdown as lifted when the first reopening order is issued. Since our focus is to explore the space of plausible lockdowns and the observed lockdowns in US states exhibit relatively little variation, we do not draw from the empirical distribution of lockdown parameters. Instead, we generate "plausible" lockdowns by drawing the duration, timing, and severity as described below.

- Duration of lockdowns: We draw the lockdown duration from a truncated exponential distribution calibrated to match the observed distribution of lockdowns in US states: the lower bound is the minimum observed time until a state reopened, the upper bound is the maximum observed time until a state reopened, and the mean is the average time to reopening.

- Timing of lockdowns: We assume the probability a policymaker implements a lockdown at time $t$ is proportional to the number of cases projected at time $t$ by a standard SIR model with no adaptive economic behavior. This produces a distribution of lockdown implementation dates which is more dispersed than what is observed, though the simulated mean is strikingly close to the empirical mean. The empirical mean is calculated as days from the first case recorded in the US.

- Severity of lockdowns: Since the degree to which lockdowns constrain consumption and labor supply is not observed (i.e., the degree to which consumers reduced consumption spending or labor hours supplied due to the lockdown is difficult to disentangle from adaptive responses to changing conditions), we draw *severity* from a uniform distribution over $[0,1]$, with $severity = 1$ representing no lockdown and $severity = 0$ indicating a total lockdown (no consumption or labor activity allowed).

Summary statistics of the data and the simulation parameters are shown in Table S6. Figure S4 shows the results for an ensemble of 2500 plausible lockdown against those of the self and targeted model outcomes. More intense lockdown (where "intensity" is the product of lockdown severity and lockdown duration) produce lower total case rates, but at a high economic cost. Across all simulated lockdown scenarios, blanket lockdown informed by a mechanistic SIR model can produce fewer cases than decentralized or coordinated behavior, though the total cost in nearly all cases exceeds the cost of the decentralized outcome.

To provide a comparison with a single blanket lockdown with voluntary and targeted isolation policies in the main text, we select the blanket lockdown with the lowest number of cases. This is consistent with the logic that the main goal of blanket lockdown is to "flatten the curve", shifting cases forward through time and reducing the number of cases overall. However, we note that the qualitative results hold for all lockdown we draw—blanket lockdown produce slightly fewer cases than voluntary isolation (and targeted isolation, for a small subset of the most severe blanket lockdown) but produce a larger economic cost (compared to targeted isolation, a vastly larger economic cost).

**Table S6:** Summary statistics for lockdown parameters

| Variable | N | Mean | St. Dev. | Min | Max |
|---|---|---|---|---|---|
| empirical duration | 51 | 48.686 | 9.501 | 31 | 77 |
| simulated duration | 2,500 | 50.396 | 13.024 | 31 | 77 |
| empirical timing | 51 | 57.137 | 2.967 | 55 | 74 |
| simulated timing | 2,500 | 57.292 | 8.931 | 23 | 98 |
| simulated severity | 2,500 | 0.505 | 0.285 | 0.001 | 0.999 |

Overall, we find blanket lockdowns tend to induce a disease-economy trade-off. This trade-off is visible in the relationship between disease and economic outcomes traced out by the simulated lockdowns. Voluntary isolation is at one end of this trade-off, favoring the economy. Targeted isolation outperforms many blanket lockdowns in terms of both disease and economic outcomes.

#### 4.2 Sensitivity to average case fatality rate (CFR)

In the remaining subsections, we describe tests of the sensitivity of our results on the difference between targeted and decentralized cases to various parameter assumptions. We find that none substantively alter our main conclusions. The results for all of the tests described in this and the following subsections are shown in Figure S5. Note that in the figure all results are presented as

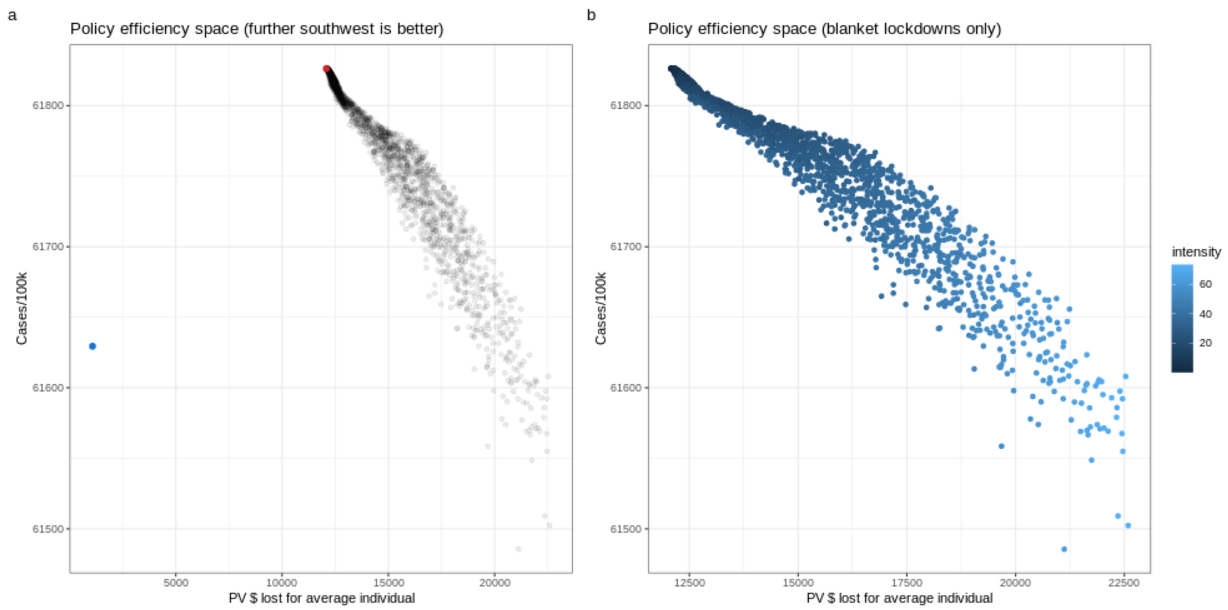

**Figure S4:** Sensitivity analysis over plausible blanket lockdown scenarios. In panel a, the red dot shows the voluntary isolation outcome while the blue dot shows the targeted isolation outcome. "intensity" in panel b is the product of *severity* and *duration*.

percentages of the measure in the targeted isolation case relative to the voluntary isolation case case (e.g. for disease outcomes, a value in the horizontal axis of 0.5 would indicate total cases in the targeted isolation case are half of the voluntary isolation case; for economic outcomes, a value of 0.8 would indicate that the targeted isolation case avoids 80% of the economic cost of the voluntary isolation case).

To study the effects of different age distributions on our findings regarding economic savings, we simulate the model at different case fatality rates covering the range of plausible values from [11]. The lower end of the range of CFRs reflects a society with a mean age near 30, while the upper end reflects a society with mean age over 80. The baseline CFR of 0.015 reflects a society with mean age between 50-59.

In general, we find that higher case fatality rates reduce the economic savings from implementing targeted isolation policies. Intuitively, this is because more severe infections produce greater permanent consequences, inducing both the planner and the decentralized individuals to avert contacts much more drastically. Averting these additional contacts is relatively costlier for the planner, because under targeted isolation the additional isolation must begin to draw from the pool of susceptibles and incur a greater economic cost.

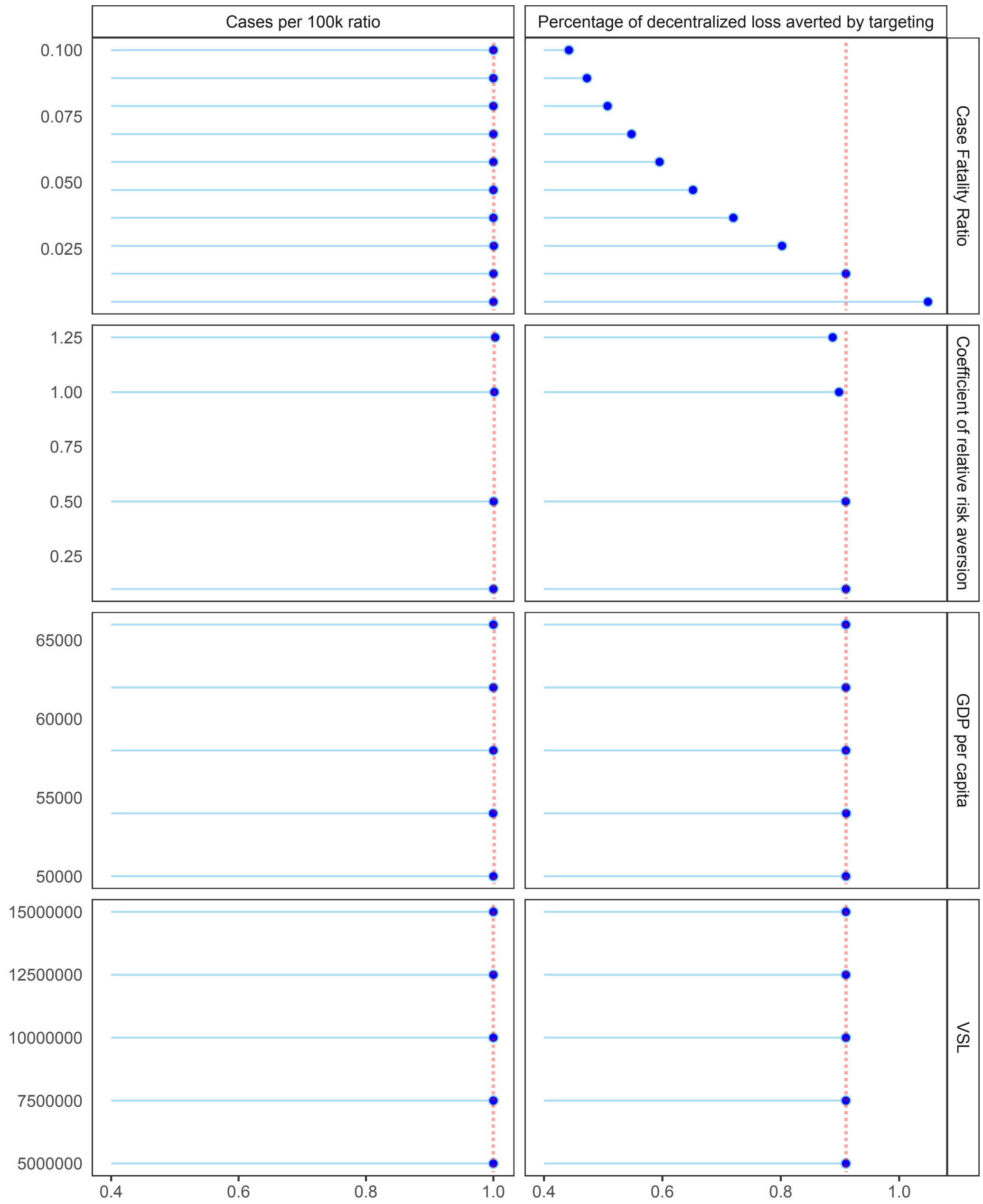

**Figure S5:** Sensitivity analyses of difference between targeted and voluntary isolation (each 2-box row relates to a separate sensitivity; each sensitivity has the labelled vertical axis). For the horizontal axes, each sensitivity shows (a) left column boxes: "Cases per 100,000" ratio (i.e. "Cases per 100,000" in the targeted isolation case divided by that same measure in the voluntary isolation case); and (b) right column boxes: percentage of decentralized loss averted by targeting (i.e. economic losses in the targeted isolation case divided by economic loss in the voluntary isolation case, then 1 minus this value). **Top row:** Plausible case fatality rates reflecting different population compositions—the dashed line shows the averted loss under the baseline model, where the CFR is taken to be 1.5%.; **Second row:** Sensitivity analysis over coefficient of relative risk aversion, $\eta$, adjusting the utility associated with death to hold the VSL constant—the dashed line shows the averted loss under the baseline model, where $\eta$ is taken to be 0.1; **Third row:** Over GDP per capita influencing daily income and therefore the cost of not working—the dashed line shows the averted loss under baseline GDP per capita of $ 58,000; **Bottom row:** Sensitivity analysis over the Value of a Statistical Life, adjusting the utility associated with death to hold the CRRA constant—the dashed line shows the averted loss under the baseline model, where VSL is $ 10m.

#### 4.3 Sensitivity to income per capita

In this section we test whether our main conclusions are sensitive to GDP/income per capita changes. The income per capita affects the daily income an individual in the model stands to make from working, as well as how much the individual would therefore have available for consumption. In this sense it captures what economic gains an individual can obtain from ignoring the disease and engaging in economic activities. It is therefore central to the overall relationship between the economy and disease.

The figure shows that income per capita does not appreciably affect the ratios – both economy and disease ratios barely move over the realistic range we test. However, income per capita does affect the overall magnitudes for both targeted isolation and voluntary isolation strategies as would be expected given the importance of this parameter for the relationship between the economy and disease.

#### 4.4 Sensitivity to VSL

Variation over this range does not appreciably affect the ratios. It does affect the magnitudes of economic losses under both planner and decentralized. We show in the next subsection (section 4.5) that variation in VSL over a much larger range (implied by holding $\Omega$ constant while altering $\eta$) does induce some change in the ratios, but is still small.

#### 4.5 Sensitivity to risk aversion

To study the sensitivity to different levels of risk aversion, we varied the coefficient of relative risk aversion ($\eta$) while holding either the utility associated with death ($\Omega$) or the Value of a Statistical Life constant. Holding either constant while altering $\eta$ implies adjustment in the other. We consider four values of $\eta$: 0.1 (used in the main model simulations), 0.5, 1 (a standard value used in financial contexts), and 1.25 (the upper bound calculated in [1]).

**Holding the VSL constant:** In this case, the utility associated with death adjusts when solving equation 11 to ensure that the trade-off between consumption and risk of death local to the pre-epidemic equilibrium reflects a VSL of $10m USD. At higher levels of risk aversion, as the risk of infection rises the planner increases the degree of isolation prescribed for susceptible individuals to reflect their consumption/risk preferences. This results in slightly lower individual recessionary savings under targeted isolation. However, the decentralized individuals do not appreciably change their behavior, since the burden of isolation was already being pushed onto the susceptible individuals. Increases in risk aversion produce no change in the decentralized behavior of recovered individuals (who face no risk) or infected individuals (who face no incentive to isolate in the absence of policy). Figure S5 summarizes these results.

**Holding the utility of death constant:** When holding the utility associated with death ($\Omega$) constant, more risk-averse individuals require greater compensation to accept the same additional risk. This additional compensation implies a higher VSL. The higher VSL induces the planner to isolate some susceptible individuals as the infection peaks to further reduce deaths, incurring greater economic costs and reducing the average individual recessionary savings under targeted isolation strategies. We show this effect by setting $\Omega = 0$ and varying $\eta$ over $(0.1, 0.5, 1, 1.25)$.[6] These values of $\eta$ imply VSLs of around \$2.7m, \$6.2m, \$174.5m, and over \$192b. Note that the total case ratio under targeted isolation relative to voluntary isolation remains unchanged—under voluntary isolation strategies, the additional adjustment is borne by the same number of additional susceptible individuals, producing the same reduction in cases and deaths.

### 5 Discussion

#### 5.1 Implications for vaccine delivery

While we do not explicitly consider vaccines or vaccine targeting, the planner's solution here provides a starting point for thinking about the rollout of vaccines.[7] Our model implies that as more individuals are vaccinated, in the decentralized equilibrium susceptible individuals will supply more labor and demand more consumption (i.e., engage in more economic activity). To the extent that vaccination reduces the effective reproductive number below 1, this vaccine-induced activity will occur until the effective reproductive number is returned to 1 (following the logic described in [24]). While maintaining $R \approx 1$ will imply similar disease dynamics as in the non-vaccine setting—a relative increase in disease prevalence relative to a non-behavioral model of vaccine delivery—it is possible that vaccination patterns will alter the burden of disease in ways our model cannot describe (e.g., [25]).

The planner would behave similarly to the decentralized equilibrium in allowing more economic activity as vaccination rates increase, but focusing on infected rather than susceptible individuals. That is, since the planner's targeted isolation strategy shields susceptible individual by isolating infected individuals, increasing rates of vaccination would enable the planner to allow

[6] The utility assocaited with death required to produce a VSL of \$10m when $\eta = 0.1$ produces even higher VSLs when $\eta > 0.1$.

[7] Incorporating vaccine targeting explicitly would require additional individual heterogeneity. In particular, it would require at least two types of susceptible individuals with different health or contact parameters.

asymptomatic infected individuals to supply labor and consume more such that $R \approx 1$ again. As in the voluntary isolation case, it is possible that such actions may worsen disease-related inequities in ways which are beyond our model's scope.

In general, the optimal vaccine rollout plan in models like ours will depend critically on both the nature of the heterogeneity in susceptible individuals as well as the objective function being maximized. For models where the social planner's objective function is an aggregation of individuals' objectives, it is highly likely that the vaccine targeting plan will involve maximizing the economic value generated given a disease progression consistent with $R \approx 1$. Thus, if the heterogeneity involves two types of susceptible individuals, for example those with high contact rates and those with low contact rates, a social planner like ours will likely concentrate vaccination efforts on those with high contact rates, as this will enable additional economic activity per vaccine delivered.

It is important to emphasize that there are multiple dimensions to vaccination targeting. As noted in [25], prioritizing those with high risk versus those with high contacts will induce different mortality/morbidity tradeoffs. These dimensions are somewhat independent of the logic of vaccinating to maximize the economic value of activity, in that they will only affect the targeting plan of a social planner like ours through how individuals value morbidity and mortality. Of course, policymakers may have different objectives. Regardless of the objective being maximized, further research with behavioral heterogeneous-agent models is urgently needed to shed light on these dimensions and potential tradeoffs.

#### 5.2 Implied hospitalization rate

We can calculate the hospitalization rate implied by the infection dynamics of our model. However this is simply an illustration, not the main focus of our analysis and we do not attempt to make detailed statements about hospital burden.

Following [20], we assume the average hospital stay is 10 days long, and calibrate the rates at which individuals recover, die, or remain infected without hospitalization to be consistent with the aggregate dynamics from the baseline model. Formally, this amounts to introducing a new "hospitalized" (H) compartment in the *SIR* model, yielding

$$S_{t+1} = S_t - \tau \mathscr{C}^{SI}(\mathbf{A}) S_t I_t, \quad (43)$$
$$I_{t+1} = \bar{P}^I I_t + \tau \mathscr{C}^{SI}(\mathbf{A}) S_t I_t - (P^H + \bar{P}^R + \bar{P}^D) I_t, \quad (44)$$
$$H_{t+1} = P^{HH} H_t + P^{IH} I_t - (P^{HR} + P^{HD}) H_t, \quad (45)$$
$$R_{t+1} = R_t + \bar{P}^R I_t + P^{HR} H_t, \quad (46)$$
$$D_{t+1} = D_t + \bar{P}^D I_t + P^{HD} H_t, \quad (47)$$

where $P^{IH} = 0.199$, $P^{HH} = 1/10$, $P^{HR} = (1 - P^{HH}) * P^{D*}$, $P^{HD} = (1 - P^{HH}) * (1 - P^{D*})$, and $(\bar{P}^I, \bar{P}^R, \bar{P}^D) = (0.784, 0.017, 0.00025)$ are calibrated to be consistent with the aggregate dynamics. Following [11], we take the baseline case fatality rate ($P^{D*}$) as 0.015. The percentage of cases hospitalized ($P^{IH}$) is calculated from a World Health Organization report measuring the percentage of cases that are severe or critical.

Figure S6 shows the results of this illustration exercise. We project that the maximum hospital burden will increase with the share of unavoidable contacts, consistent with evidence regarding household transmission [26, 11]. Similarly, we project that the maximum hospital burden will be highest when consumption contacts are low relative to labor contacts. In such cases, individuals face lower incentives to reduce economic activities and voluntarily isolate, generating higher infection profiles as a result.

The hospitalization rates implied by our model are consistent with estimates from the literature, e.g., [11]. We do not model the costs of approaching or exceeding hospital capacity. Including a convex cost for exceeding the hospital capacity constraint would induce the planner to maintain lower infection rates, with the magnitude of the additional infection avoidance depending on the value of the penalty [27]. What is less clear is how such a penalty would change decentralized behavior. This will depend on the nature of healthcare markets and how effectively crowding costs at hospitals are channeled to individuals, and the degree to which individuals are able to anticipate these crowding costs and act accordingly. It is not obvious what proportion of these costs are or should be internalized by decentralized individuals.

#### 5.3 Implications of blanket lockdowns on hospitalization rates

While our analysis suggests that blanket lockdowns are generally costlier in economic terms than voluntary isolation under universal testing, ignoring the costs of exceeding hospital capacity [11] ignores a major channel for potential savings from blanket lockdowns. If blanket lockdowns are

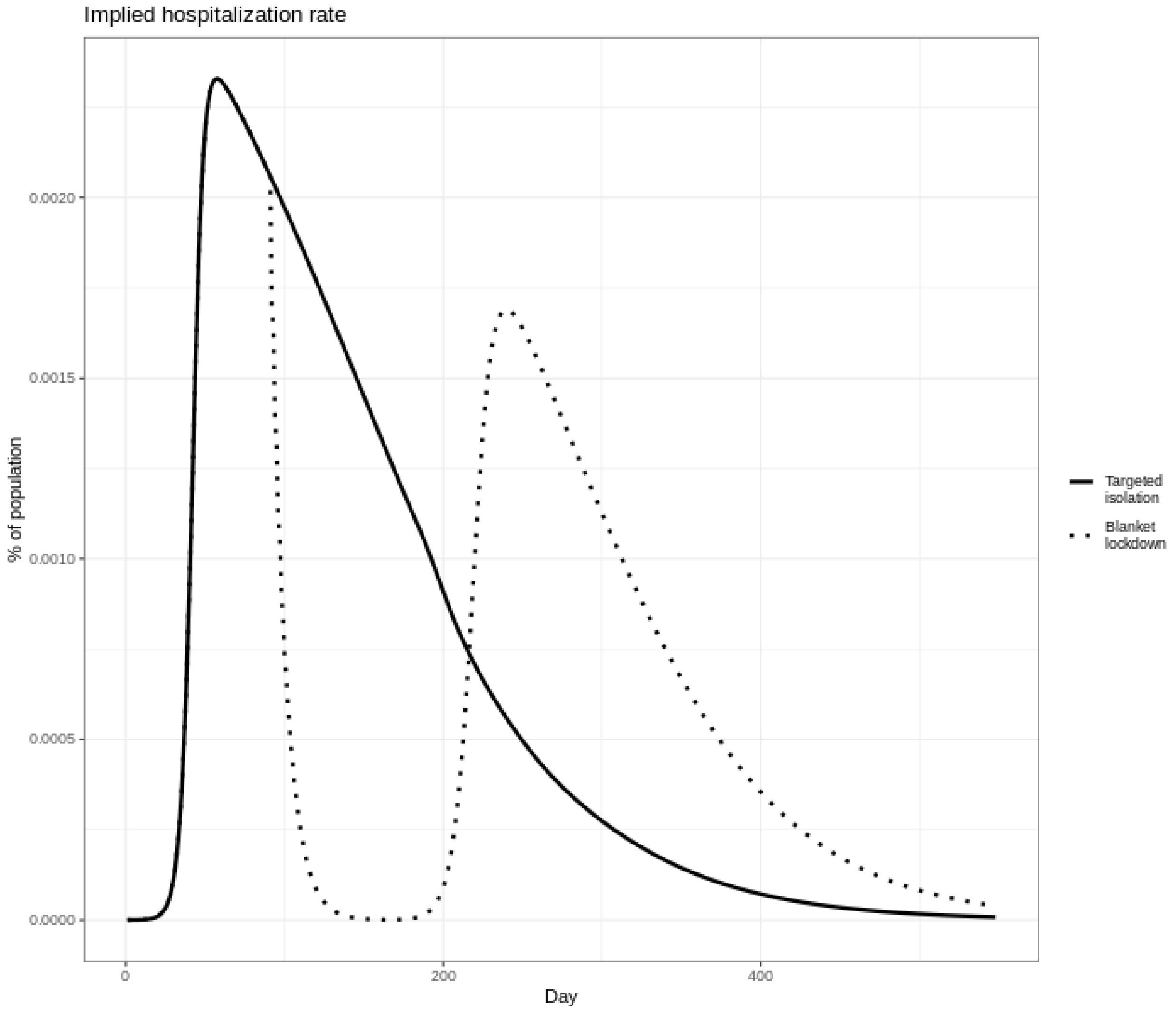

**Figure S6:** Implied hospitalization rate under targeted isolation and the case-minimizing blanket lockdown shown in Figure 2 of the main text.

able to shift infections forward to periods when there is lower strain on the healthcare system, then blanket lockdowns may generate nontrivial savings relative to voluntary isolation under universal testing. Figure S7 shows the time paths of GDP deviation and infection prevalence under the ensemble of plausible blanket lockdowns we simulate (see section 4.1), along with the associated marginal distributions of total economic losses and cases per 100,000.

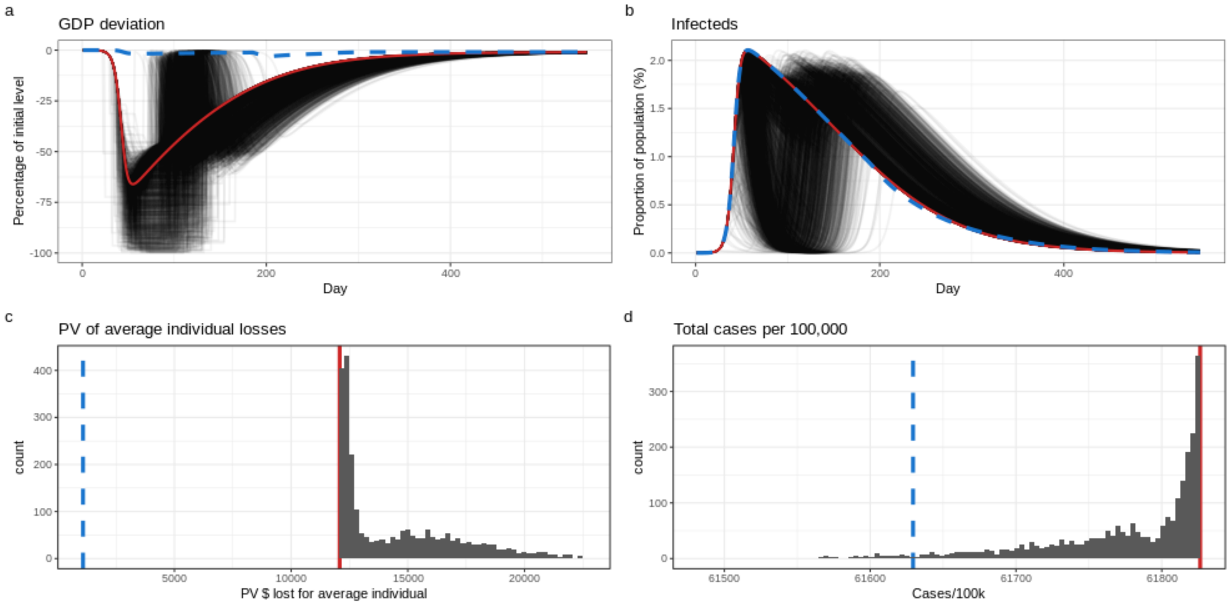

**Figure S7:** Sensitivity analysis over plausible blanket lockdown scenarios. In panels a and b, the solid red line shows the voluntary isolation path and the dashed blue line shows the targeted isolation path, while the light black lines show alternative blanket lockdown scenarios. In panels c and d, the solid red line shows the voluntary isolation outcome while the dashed blue line shows the targeted isolation outcome.

We observe that lockdowns which begin earlier and last longer lead to lower peak infection rates. However, offsetting this potential benefit, relaxing blanket lockdowns tends to cause rebound effects. At the end of a blanket lockdown, susceptible individuals take advantage of the lower risk of infection and engage in contactful activities, causing a spike in cases. Such rebound effects are observed in empirical data from COVID-19 as well as in other environmental settings where policies induce individuals to shift behavior through time (e.g. investment rebounds where policy uncertainty is resolved, pent-up demand, etc.). However, if the lockdown lasts long enough, there will be few enough infected individuals left that the rebound effect will produce a lower peak. These results suggest that blanket lockdowns aimed at preventing hospital capacity from being overwhelmed must be used with caution. The blanket lockdown must begin early enough to prevent the no-intervention peak in cases, and must last long enough that the rebound effect will not produce a similar peak.

#### 5.4 Additional considerations

Below we list and briefly describe some additional considerations in developing, interpreting, and extending coupled-systems models such as ours.

##### 5.4.1 Cross-regional comparisons

The importance of model calibration points to the fact that cross-regional (e.g. inter-state or inter-country) comparisons need to be made "ceteris paribus". That is, the model has a range of parameters that represent a given population. These parameters will be different across different countries and states, meaning that states should be judged against a (potentially counterfactual) state with the same parameterization but a different policy approach. Comparing results of policy outcomes across regions with different parameterizations will result in inferences which mix the effects of the policy approaches with the effects of different parameter values.

##### 5.4.2 Different economic sectors

The possibility of remote work varies by sector. Workers without this option face a choice between whether they go into work and risk contracting the infection or stay at home and lose out financially from being unable to work. Our contact function reflects this dynamic: labor supply is associated with positive contact levels. A smaller labor coefficient in our contact function would imply workers could continue working without many contacts, reflecting the ability to work from home. By contrast a high labor coefficient would imply changing hours worked causes contacts to change a lot, reflecting less ability to work from home. This same approach can be taken to modelling contactless consumption, where low contact coefficients on consumption reflect the ability to consume contactlessly (i.e. changes in consumption demand have a smaller effect on contacts). Figure 4 in the main text shows our main result is qualitatively unchanged when these coefficients are adjusted.

##### 5.4.3 Behavioral adjustment costs

Individuals in the real world often take time to adjust their behavior in response to changing conditions. Whatever the reason, such adjustment costs might shift the dynamics of infection in a small time horizon until individuals' behavior is adjusted. However, adjustment costs would not qualitatively change the dynamics of the model or the rationale underlying the coordination failure, since individuals would eventually make the adjustments.

##### 5.4.4 Alternative social welfare functions

The targeted isolation strategy is the solution to an optimization problem where individual consumption and labor supply by each health type is chosen to maximize a population-weighted sum of utilities of each health type. Alternative social welfare functions may produce different strategies. For example, a different weighting scheme for type-specific utilities may alter the degree of targeted isolation.

We examined the solution to the unweighted problem (i.e. using a social welfare function with type-specific utilities unweighted by population size, implying all infected individuals are given equal consideration as all susceptible individuals) and found that the targeted isolation strategy was still followed, indicating that the welfare losses to the average susceptible individual from reducing activity outweighed the welfare losses to the average infected individual. Alternative social welfare functions, such as a purely-mortality focused objective, may produce substantially different strategies (e.g. a welfare function which considered only total deaths may produce a strategy which suppresses the infection immediately despite a large economic cost) but are beyond our scope here. Fundamentally, any objective function implies some weighting over the interests of individuals with different health types, and any policy involves trading off the interests of these different groups. Population weighting implies that each individual's interests are considered equally regardless of health type.

#### References

[1] Raj Chetty. Consumption commitments, unemployment durations, and local risk aversion. Technical report, National Bureau of Economic Research, 2004.

[2] Robert McClelland and Shannon Mok. A review of recent research on labor supply elasticities. Technical report, 2012.

[8] US Environmental Protection Agency. Mortality risk valuation: What value of statistical life does epa use? Technical report, 2020.

[9] Martin S Eichenbaum, Sergio Rebelo, and Mathias Trabandt. The macroeconomics of epidemics. Working Paper 26882, National Bureau of Economic Research, 2020.

[10] Juanjuan Zhang, Maria Litvinova, Wei Wang, Yan Wang, Xiaowei Deng, Xinghui Chen, Mei Li, Wen Zheng, Lan Yi, Xinhua Chen, Qianhui Wu, Yuxia Liang, Xiling Wang, Juan Yang, Kaiyuan Sun, Ira M. Longini, M. Elizabeth Halloran, Peng Wu, Benjamin J. Cowling, Stefano Merler, Cecile Viboud, Alessandro Vespignani, Marco Ajelli, and Hongjie Yu. Evolving epidemiology and transmission dynamics of coronavirus disease 2019 outside Hubei province, China: a descriptive and modelling study. *The Lancet Infectious Diseases*, 20(7):793–802, July 2020.

[11] Robert Verity, Lucy C Okell, Ilaria Dorigatti, Peter Winskill, Charles Whittaker, Natsuko Imai, Gina Cuomo-Dannenburg, Hayley Thompson, Patrick GT Walker, Han Fu, et al. Estimates of the severity of coronavirus disease 2019: a model-based analysis. *The Lancet infectious diseases*, 2020.

[12] The Centre for Evidence-Based Medicine. Global covid-19 case fatality rates. https://www.cebm.net/covid-19/global-covid-19-case-fatality-rates/.

[13] Kiesha Prem, Alex Cook, and Mark Jit. Projecting social contact matrices in 152 countries using contact surveys and demographic data. *PLOS Computational Biology*, 13(9), 2017.

[14] O Diekmann, JAP Heesterbeek, and MG Roberts. The construction of next-generation matrices for compartmental epidemic models. *Journal of the Royal Society, Interface*, 7(47):873–885, 2010.

[15] Robert E Lucas Jr. Econometric policy evaluation: A critique. In *Carnegie-Rochester con-*

*ference series on public policy*, volume 1, pages 19–46. North-Holland, 1976.

[16] Christopher I Jarvis, Kevin Van Zandvoort, Amy Gimma, Kiesha Prem, Petra Klepac, G James Rubin, and W John Edmunds. Quantifying the impact of physical distance measures on the transmission of covid-19 in the uk. *BMC medicine*, 18:1–10, 2020.

[17] Benjamin M Althouse, Edward A Wenger, Joel C Miller, Samuel V Scarpino, Antoine Allard, Laurent Hébert-Dufresne, and Hao Hu. Stochasticity and heterogeneity in the transmission dynamics of sars-cov-2. *arXiv preprint arXiv:2005.13689*, 2020.

[18] Lawrence H Goulder, Marc AC Hafstead, GyuRim Kim, and Xianling Long. Impacts of a carbon tax across us household income groups: What are the equity-efficiency trade-offs? *Journal of Public Economics*, 175:44–64, 2019.

[19] Bureau of Labor Statistics. American time use, 2019. Data retrieved from BLS website, series TUU10101AA01002867 https://data.bls.gov/timeseries/TUU10101AA01002867.

[20] Dawei Wang, Bo Hu, Chang Hu, Fangfang Zhu, Xing Liu, Jing Zhang, Binbin Wang, Hui Xiang, Zhenshun Cheng, Yong Xiong, Yan Zhao, Yirong Li, Xinghuan Wang, and Zhiyong Peng. Clinical Characteristics of 138 Hospitalized Patients With 2019 Novel Coronavirus–Infected Pneumonia in Wuhan, China. *Journal of the American Medical Association*, 323(11):1061–1069, 03 2020.

[21] World Health Organization. Report of the who–china joint mission on coronavirus disease 2019 (covid-19). Technical report, 2020.

[22] Yongyang Cai and Kenneth L Judd. Stable and efficient computational methods for dynamic programming. *Journal of the European Economic Association*, 8(2-3):626–634, 2010.

[23] Eli P Fenichel, Carlos Castillo-Chavez, M Graziano Ceddia, Gerardo Chowell, Paula A Gonzalez Parra, Graham J Hickling, Garth Holloway, Richard Horan, Benjamin Morin, Charles Perrings, et al. Adaptive human behavior in epidemiological models. *Proceedings of the National Academy of Sciences*, 108(15):6306–6311, 2011.

[24] Joshua S Gans. The economic consequences of r = 1: Towards a workable behavioural epidemiological model of pandemics. Working Paper 27632, National Bureau of Economic Research, July 2020.

[25] Shweta Bansal, Babak Pourbohloul, and Lauren Ancel Meyers. A comparative analysis of influenza vaccination programs. *PLoS Med*, 3(10):e387, 2006.

[26] Kiesha Prem, Yang Liu, Timothy W Russell, Adam J Kucharski, Rosalind M Eggo, Nicholas Davies, Stefan Flasche, Samuel Clifford, Carl AB Pearson, James D Munday, et al. The effect of control strategies to reduce social mixing on outcomes of the covid-19 epidemic in wuhan, china: a modelling study. *The Lancet Public Health*, 2020.

[27] Daron Acemoglu, Victor Chernozhukov, Iván Werning, and Michael D Whinston. A multi-risk sir model with optimally targeted lockdown. Working Paper 27102, National Bureau of Economic Research, 2020.
